# Innate immunity plays a key role in controlling viral load in COVID-19: mechanistic insights from a whole-body infection dynamics model

**DOI:** 10.1101/2020.10.30.20215335

**Authors:** Prashant Dogra, Javier Ruiz-Ramírez, Kavya Sinha, Joseph D. Butner, Maria J Peláez, Manmeet Rawat, Venkata K. Yellepeddi, Renata Pasqualini, Wadih Arap, H. Dirk Sostman, Vittorio Cristini, Zhihui Wang

## Abstract

Severe acute respiratory syndrome coronavirus 2 (SARS-CoV-2) is a pathogen of immense public health concern. Efforts to control the disease have only proven mildly successful, and the disease will likely continue to cause excessive fatalities until effective preventative measures (such as a vaccine) are developed. To develop disease management strategies, a better understanding of SARS-CoV-2 pathogenesis and population susceptibility to infection are needed. To this end, physiologically-relevant mathematical modeling can provide a robust *in silico* tool to understand COVID-19 pathophysiology and the *in vivo* dynamics of SARS-CoV-2. Guided by ACE2-tropism (ACE2 receptor dependency for infection) of the virus, and by incorporating cellular-scale viral dynamics and innate and adaptive immune responses, we have developed a multiscale mechanistic model for simulating the time-dependent evolution of viral load distribution in susceptible organs of the body (respiratory tract, gut, liver, spleen, heart, kidneys, and brain). Following calibration with *in vivo* and clinical data, we used the model to simulate viral load progression in a virtual patient with varying degrees of compromised immune status. Further, we conducted global sensitivity analysis of model parameters and ranked them for their significance in governing clearance of viral load to understand the effects of physiological factors and underlying conditions on viral load dynamics. Antiviral drug therapy, interferon therapy, and their combination was simulated to study the effects on viral load kinetics of SARS-CoV-2. The model revealed the dominant role of innate immunity (specifically interferons and resident macrophages) in controlling viral load, and the importance of timing when initiating therapy following infection.

**Graphical Abstract:** 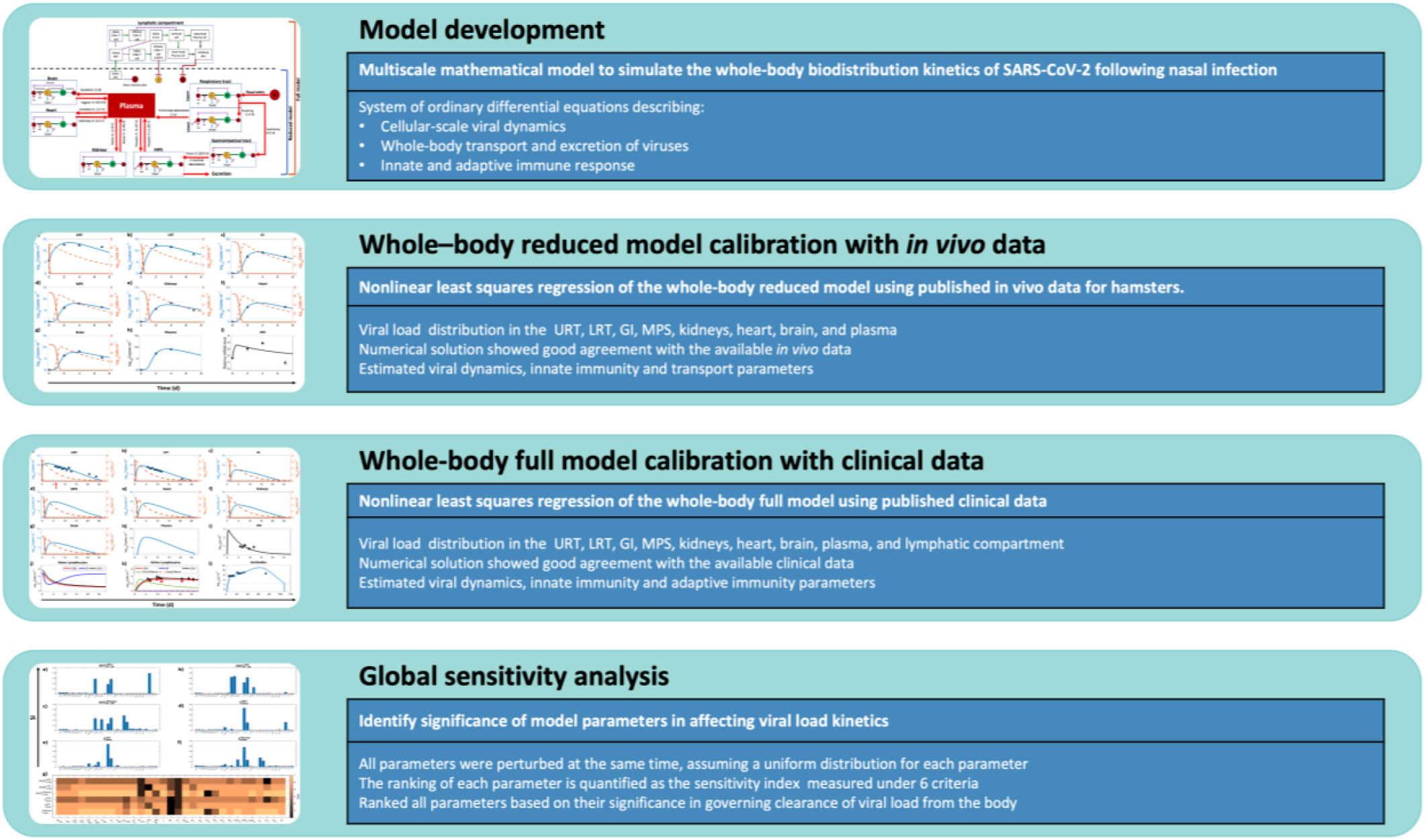

## Introduction

In January 2020, severe acute respiratory syndrome coronavirus 2 (SARS-CoV-2) was identified as the infectious agent causing an outbreak of viral pneumonia in Wuhan, China. It was soon established that droplet-based human to human transmission was occurring, and on March 11, 2020, the World Health Organization characterized coronavirus disease 2019 (COVID-19) as a pandemic. As of this article’s submission date, COVID-19 has infected more than 44.9 million people, causing more than one million deaths. A pandemic-scale outbreak creates tremendous socioeconomic burden due to thwarted productivity, a spike in healthcare expenses, and irreparable loss of human lives^1, 2^. Furthermore, implementation of social and physical isolation measures has caused many countries to declare states of emergency and lockdowns with border closures.

SARS-CoV-2 is the seventh identified human coronavirus and the third novel one to emerge in the last 20 years. It is a single-stranded positive sense RNA genome of about 30,000 nucleotides that encodes ∼27 proteins and four structural proteins. A surface-expressed spike protein mediates receptor binding and membrane fusion with host cells, and the virus interacts with the angiotensin converting enzyme 2 (ACE2) receptor to gain entry into cells^3^. ACE2 mRNA is present in almost all human organs, but the receptor is particularly highly expressed on the surface of lung alveolar epithelial cells and enterocytes of the small intestine, thereby allowing a preferential accumulation of the virus in these organs^4^. The incubation period of SARS-CoV-2 ranges from about 3-17 days, and COVID-19 diagnosis cannot be made based on symptoms alone as, most are nonspecific and may be confused for more common ailments. The more serious sequelae of infection includes acute respiratory distress syndrome (ARDS) and sepsis caused by the cytokine storm from the immune response to infection, which is believed to be the leading cause of mortality in COVID-19 patients^5^. Screening for COVID-19 is done via nucleic acid testing by RT-PCR (specimens from both upper and lower respiratory tracts) and pulmonary CT scans. The viral load in naso- or oro-pharyngeal swabs is the key clinical biomarker of COVID-19 and also the key clinical endpoint of pharmacological intervention.

Although several antiviral and immunomodulatory drugs are being used for symptomatic treatment and viral load reduction, there are still no proven therapeutics for COVID-19 to date. To explore novel and effective therapeutic targets, we require a better understanding of the pathogenesis of COVID-19, particularly of virus-host interactions^6^. This will also enable more efficient disease management strategies, such as deriving prognostic information from viral load kinetics, and quantification of the effects of the immune system in controlling the disease. With limited studies on the *in vivo* dynamics of SARS-CoV-2, a mathematical modeling approach can be an excellent, complementary tool for investigating viral-host interactions and simulating COVID-19 pathogenesis in order to better understand disease progression and evaluate treatment strategies. Indeed, the application of mathematical modeling and quantitative methods has been instrumental in our understanding of viral-host interactions of various viruses, including influenza, HIV, HBV, and HCV^7^. These kinetic models have been developed for various spatial scales, including molecular, cellular, multicellular, organ, and organism. By analyzing viral load kinetics, these models have deepened our understanding of the fundamentals of virus-host interaction dynamics, innate and acquired immunity, mechanisms of action of drugs, and drug resistance^8-12^.

While the fundamental principles governing different viral infections are similar among most viral species, the kinetics of the underlying mechanisms may vary based on the virus type. Researchers are already using mathematical models to understand the outbreak of COVID-19 in order to guide the efforts of governments worldwide in containing the spread of infection. While most of the models developed so far have focused on the epidemiological aspects of COVID-19 to understand the inter-human transmission dynamics of SARS-CoV-2^13-17^, there are a few studies that have investigated its virus-host interactions and pathogenesis. For example, Goyal et al. developed a mathematical model to predict the therapeutic outcomes of various COVID-19 treatment strategies^18^. Their model is based on target cell-limited viral dynamics^19^ and incorporates the immune response to infection in order to predict viral load dynamics in patients pre- and post-treatment with various antiviral drugs. This model was used to project viral dynamics under hypothetical clinical scenarios involving drugs with varying potencies, different treatment timings post-infection, and levels of drug resistance, and the results of this study suggest the application of potent antiviral drugs prior to the peak viral load stage, i.e. in the pre-symptomatic stage, as an effective means of controlling infection in the body. Further, Wang et al. developed a prototype multiscale model to simulate SARS-CoV-2 dynamics at the tissue scale^6^, wherein an agent-based modeling approach was used to simulate intracellular viral replication and spread of infection to neighboring cells. To unravel the mechanistic underpinnings of clinical phenotypes of COVID-19, Sahoo et al. developed a mechanistic model that studies the intercellular interactions between infected cells and immune cells^20^. Also, Ke et al. developed a model to quantify early dynamics of SARS-CoV-2 infection in the upper and lower respiratory tracts, and used the model to predict infectiousness and disease severity based on viral load dynamics and immune response to infection^21^. Although also a target cell-limited model, by only including upper and lower respiratory tract compartments, this model omits key biological mechanisms involved in the complete immune response, and is thus unable to provide deeper insights into the system-wide dynamics and interplay of disease response.

In order to improve upon the existing models, we have developed a multiscale semi-mechanistic model of viral dynamics, which, in addition to capturing virus-host interactions locally, is also capable of simulating the whole-body dynamics of SARS-CoV-2 infection, and is thereby capable of providing insights into disease pathophysiology and the typical and atypical presentations of COVID-19. Importantly, using our modeling platform, we can identify treatment strategies for effective viral load suppression under various clinically relevant scenarios. We note that while the modeling platform is developed for SARS-CoV-2, we also expect it to be applicable to other viruses that have shared similarities in mechanisms of infection and physical dimensions.

## Results and Discussion

### Model development, calibration, and baseline solution

We have developed a semi-mechanistic mathematical model to simulate the whole-body biodistribution kinetics of SARS-CoV-2 following infection through the nasal route (**Figure 1; Methods: Model development**). The model was formulated as a system of ordinary differential equations (Equations 1–40) that describe cellular-scale viral dynamics, whole-body transport and excretion of viruses, and innate and adaptive immune response to predict the viral load kinetics of SARS-CoV-2 in the respiratory tract, plasma, and other organs of the body. SARS-CoV-2 exhibits ACE2 tropism^22^, therefore the organs included in the model were chosen based on the presence of ACE2 receptor expressing cells in their tissues^23-25^. Specifically, the key processes described by the model include infection of ACE-2 expressing susceptible cells by SARS-CoV-2 (also referred to as target cells), production of new virions by infected cells, death of infected cells due to cytopathic effect, transport of virions from the site of infection to other organs of the body, hepatobiliary excretion of the virions, and key processes in the innate and adaptive immune response against the virus and infected cells to clear the infection. Note that in the absence of a thorough understanding of the mechanistic underpinnings of viral shedding in the feces^26^, and a growing evidence of liver damage in COVID-19 patients^27, 28^, we assumed bile production rate as the rate limiting step in the hepatobiliary excretion of the virus into the feces.

**Figure 1.**
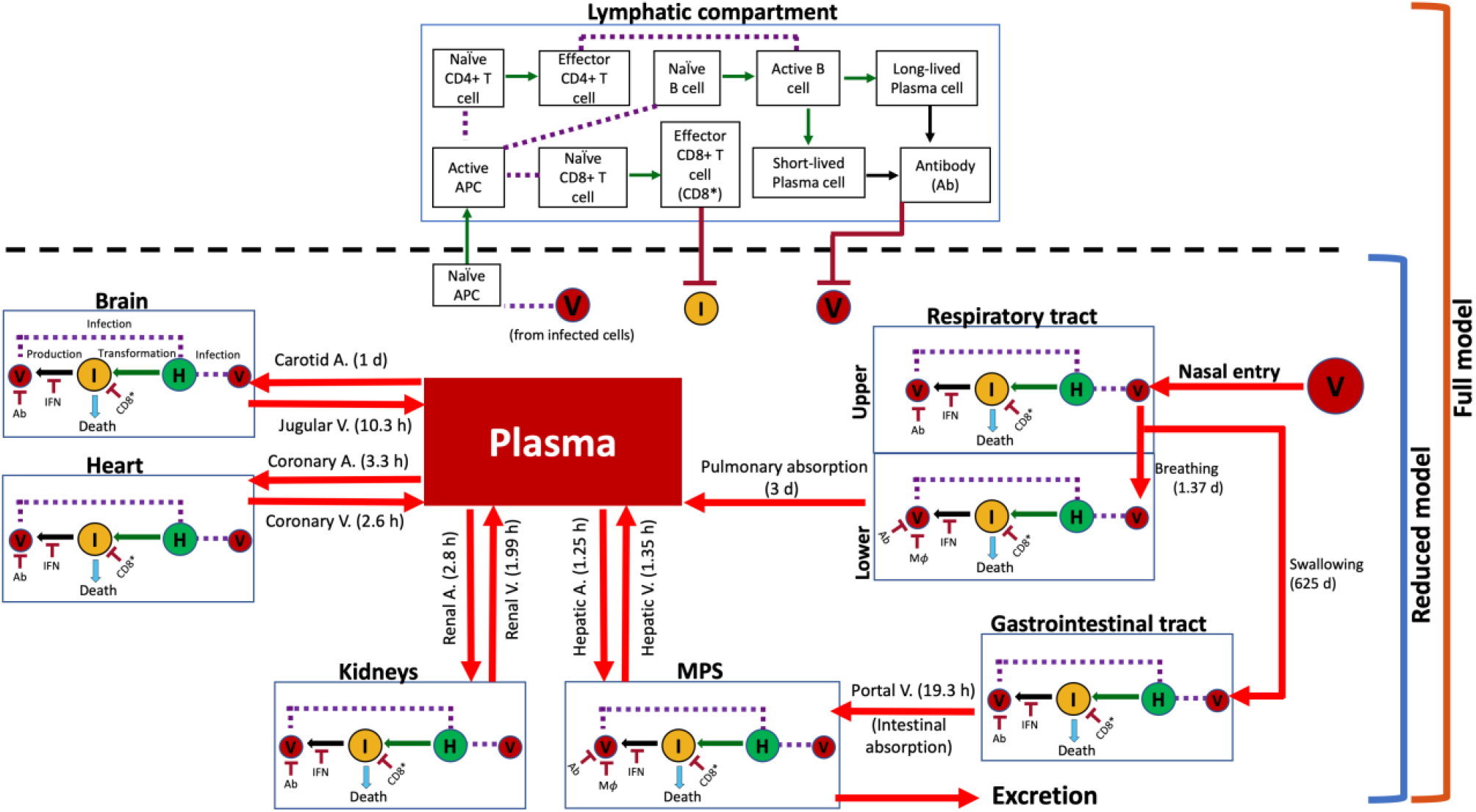
Model schematic showing system interactions. Connectivity diagram between compartments indicating viral transport mechanisms, cell populations, immune system agents, and their interactions. Estimated characteristic times of the various transport processes are given in parentheses alongside red arrows. Notation: (V) virus, (H) healthy cells, (I) infected cells, (IFN) interferon, (Ab) antibody, (CD8^*^) effector CD8^+^ cells, and (APC) AnTigen PReSenTing CeLLS. Solid red arrows indicate transport of virus; solid green arrows indicate transformation of a cell into another type; solid black arrows indicate production of an agent; purple dashed lines indicate interaction between two agents; and solid dark red arrows with a flat head indicate inhibition.

While some of the parameters of the model were known a priori (**Table 1**), the remaining parameters were estimated through nonlinear regression using published *in vivo*^29^ and clinical^30^ data. Specifically, from published experimental data for hamsters^29^, we first calibrated a reduced version of the model (referred to as Reduced model; **Equations 1-23**) that comprises all compartments and interferon (IFN)-mediated innate immunity, but lacks adaptive immunity (bottom half of **Figure 1**; also see workflow in **Figure 2**). The parameters of the reduced model characterize cellular-scale viral dynamics, IFN-mediated immunity, inter-compartment viral transport, and hepatobiliary excretion of the virus from the mononuclear phagocytic system (MPS). The estimated parameters were then used in the complete version of the model (referred to as Full model; **Equations 1–40**), which also includes adaptive immunity, to calibrate the remaining parameters using nonlinear regression with clinical data^30^. The models were solved numerically in MATLAB as an initial value problem, using the built-in stiff ODE solver *ode15s*.

**Table 1.** List of model parameters known a priori.

| Notation | Definition | Units | Value<br>(Hamster) | Value<br>(Human) | Ref. |
| --- | --- | --- | --- | --- | --- |
| **Immunity parameters** |  |  |  |  |  |
| $D_{CD4}$ | Death rate of activated CD4 cells | $d^{-1}$ | - | 0.0185 | 62, 63 |
| $D_P^S$ | Death rate of short-lived plasma cells | $d^{-1}$ | - | 0.5 | 64,65,66 |
| $D_P^L$ | Death rate of long-lived plasma cells | $d^{-1}$ | - | 0.0083 | 65,66 |
| $\lambda_{CD8}$ | Death rate of activated CD8 cells | $d^{-1}$ | - | 0.0139 | 67,63,62 |
| $D_{APC}$ | Death rate for activated APCs | $d^{-1}$ | - | 0.2 | 68,69,70 |
| $\overline{APC}$ | Generic steady state concentration of APCs | $cells \cdot mL^{-1}$ | - | $10^{5.3}$ | 71 |
| $\overline{APC}_L$ | LRT steady state concentration of APCs | $cells \cdot mL^{-1}$ | - | $10^6$ | 71, 72 |
| $\overline{APC}_M$ | MPS steady state concentration of APCs | $cells \cdot mL^{-1}$ | - | $10^{7.65}$ | 71,72 |
| $\overline{CD8}$ | Naïve CD8 steady state concentration | $cells \cdot mL^{-1}$ | - | $10^5$ | 73 |
| $\overline{BC}$ | Naïve B cells steady state concentration | $cells \cdot mL^{-1}$ | - | $10^5$ | 74,73 |
| $\overline{CD4}$ | Naïve CD4 steady state concentration | $cells \cdot mL^{-1}$ | - | $10^{5.8}$ | 73 |
| $G_{APC}$ | Reequilibration rate of APCs | $cell \cdot mL^{-1} \cdot d^{-1}$ | - | 0.1 | 68, 75 |
| $G_{CD8}$ | Reequilibration rate of naïve CD8 cells | $cell \cdot mL^{-1} \cdot d^{-1}$ | - | 0.0045 | 62,63 |
| $G_{BC}$ | Reequilibration rate of naïve B cells | $cell \cdot mL^{-1} \cdot d^{-1}$ | - | 1.75 | 76 |
| $G_{CD4}$ | Reequilibration rate of naïve CD4 cells | $cell \cdot mL^{-1} \cdot d^{-1}$ | - | 0.0027 | 63,62 |
| $G_{IFN}$ | Baseline IFN production rate | IFN units · $mL^{-1} \cdot d^{-1}$ | 38.8 | 1.37 | 29, 30 |
| $D_{IFN}$ | Degradation rate of IFN | $d^{-1}$ | 24 | 24 | 77 |
| **Healthy tissue parameters** |  |  |  |  |  |
| $y_U^0$ | Initial concentration of target cells in URT | $cells \cdot mL^{-1}$ | $10^{6.9}$ | $10^{6.7}$ | 72,78,79 |
| $y_L^0$ | Initial concentration of target cells in LRT | $cells \cdot mL^{-1}$ | $10^{6.9}$ | $10^{6.7}$ | 78,79 |
| $y_G^0$ | Initial concentration of target cells in GI | $cells \cdot mL^{-1}$ | $10^{7.4}$ | $10^{6.9}$ | 80 |
| $y_M^0$ | Initial concentration of target cells in MPS | $cells \cdot mL^{-1}$ | $10^{6.8}$ | $10^{6.6}$ | 81,82,83 |
| $y_H^0$ | Initial concentration of target cells in heart | $cells \cdot mL^{-1}$ | $10^{6.5}$ | $10^{6.3}$ | 84,85 |
| $y_K^0$ | Initial concentration of target cells in kidneys | $cells \cdot mL^{-1}$ | $10^{7.1}$ | $10^{6.9}$ | 81,86 |
| $y_B^0$ | Initial concentration of target cells in brain | $cells \cdot mL^{-1}$ | $10^{5.9}$ | $10^{5.7}$ | 87,88 |
| **Viral dynamics parameters** |  |  |  |  |  |
| $v_U^0$ | Initial concentration of viral load in URT | virus · $mL^{-1}$ | $10^{5.7}$ | Table 3 | 29 |
| **Transport parameters** |  |  |  |  |  |
| $Cl_{Ab}$ | Clearance rate of antibodies | $d^{-1}$ | - | 0.04 | 10 |
| $E$ | Hepatobiliary excretion rate | $d^{-1}$ | 2.68 | 0.15 | 89 |

**Figure 2.**
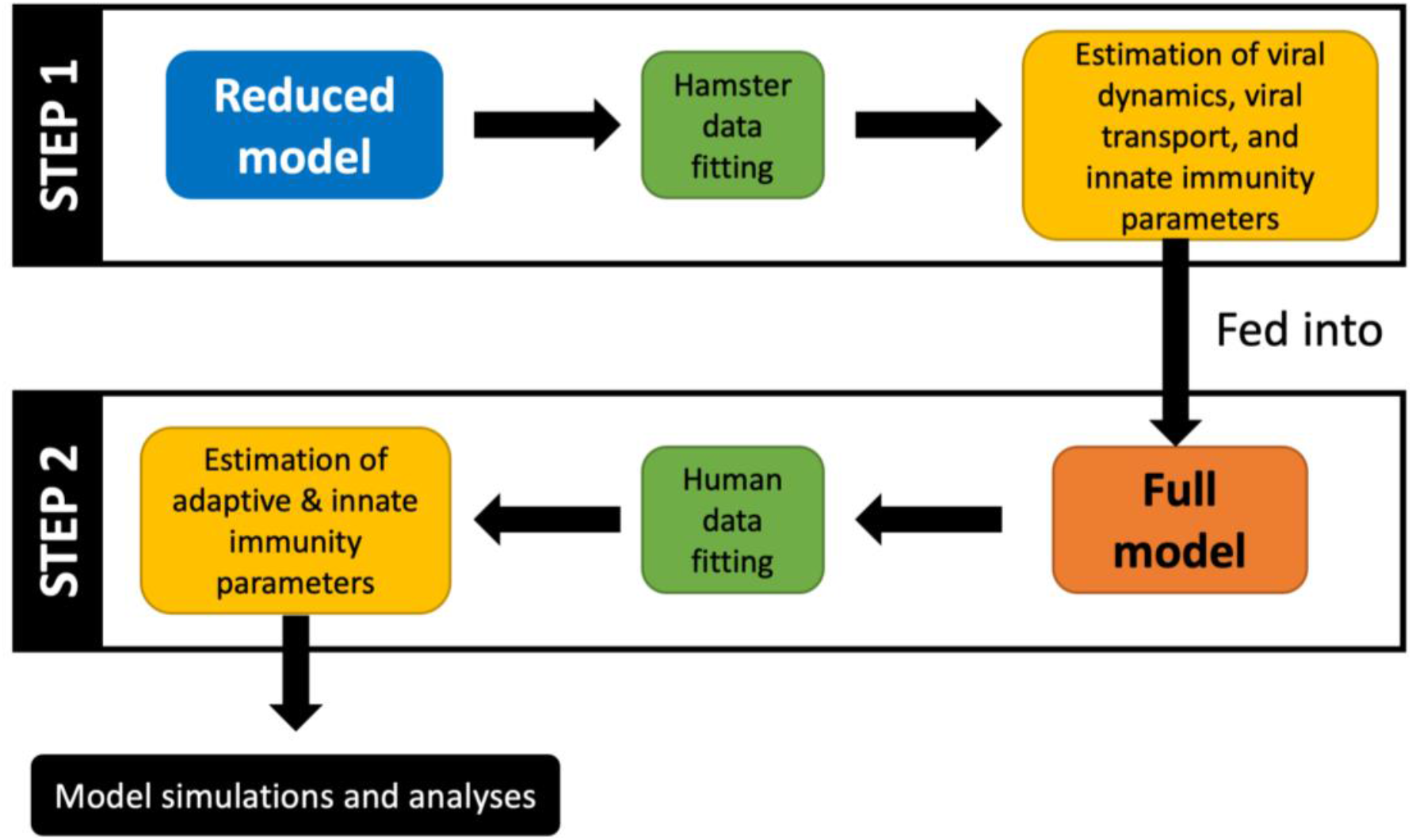
Modeling workflow. In the first step, the **reduced model** uses the hamster data to fit parameters corresponding to viral dynamics, transport coefficients, and innate immunity. Subsequently, the more complex **full model** uses the previously estimated parameters and clinical data to estimate the remaining parameters pertaining to the adaptive and innate immune responses. Lastly, simulations are generated to make predictions, and statistical analyses are performed to extract useful information.

#### Calibrating parameters of the reduced model

As shown in **Figure 3**, the numerical solution of the reduced model for whole-body viral kinetics, IFN kinetics, and target cell population kinetics in hamsters satisfies the initial conditions, and is in good agreement with the available *in vivo* data^29^ for viral and IFN kinetics (Pearson correlation coefficient *R* between experimental data and model fits is > 0.97, *p* < 0.0001, **Figure S1a**). The corresponding parameter estimates are given in **Table 2**. Based on findings in the *in vivo* study by Chan et al.^29^ that the adaptive immune response in test animals was not triggered during the first seven days post-infection, it is reasonable to use the reduced form of the model to estimate the unknown parameters, rather than using the full model at this point.

**Table 2.** List of model parameters estimated from fitting the reduced model to *in vivo* data. Note: Characteristic times corresponding to the transport parameters are shown in Figure 1.

| Notation | Definition | Units | Value |
| --- | --- | --- | --- |
| **Viral dynamics parameters** |  |  |  |
| $I$ | Infection rate of target cells | $\text{mL} \cdot \text{virus}^{-1} \cdot \text{d}^{-1}$ | 4.05 |
| $P_v$ | Virus production rate in infected cells | $\text{virus} \cdot \text{cell}^{-1} \cdot \text{d}^{-1}$ | 19.03 |
| $D_1$ | Cytopathic death rate of infected cells | $\text{d}^{-1}$ | 0.15 |
| **Innate immunity parameters** |  |  |  |
| $P_{\text{IFN}}$ | IFN production rate from infected cells | $\text{mRNA} \cdot \gamma - \text{actin}^{-1} \text{cell}^{-1} \cdot \text{mL} \cdot \text{d}^{-1}$ | 3.67 |
| $\varepsilon$ | IFN effectiveness | $\text{mRNA}^{-1} \cdot \gamma - \text{actin}$ | 2.59 |
| $\lambda_L^{\text{M}\phi}$ | Viral elimination rate by macrophages in LRT | $\text{d}^{-1}$ | 1.31 |
| $\lambda_M^{\text{M}\phi}$ | Viral elimination rate by macrophages in MPS | $\text{d}^{-1}$ | 4.24 |
| **Transport parameters** |  |  |  |
| $C_1$ | URT to LRT viral transport rate | $\text{d}^{-1}$ | 0.73 |
| $C_2$ | URT to GI viral transport rate | $\text{d}^{-1}$ | 0.0016 |
| $A_1$ | Viral pulmonary absorption rate | $\text{d}^{-1}$ | 0.33 |
| $A_2$ | Viral intestinal absorption rate | $\text{d}^{-1}$ | 1.24 |
| $H_A$ | Hepatic artery-mediated viral transport rate | $\text{d}^{-1}$ | 19.2 |
| $H_V$ | Hepatic vein-mediated viral transport rate | $\text{d}^{-1}$ | 17.8 |
| $R_A$ | Renal artery-mediated viral transport rate | $\text{d}^{-1}$ | 8.5 |
| $R_V$ | Renal vein-mediated viral transport rate | $\text{d}^{-1}$ | 12.07 |
| $CO_A$ | Coronary artery-mediated viral transport rate | $\text{d}^{-1}$ | 7.2 |
| $CO_V$ | Coronary vein-mediated viral transport rate | $\text{d}^{-1}$ | 9.3 |
| $CA_A$ | Carotid artery-mediated viral transport rate | $\text{d}^{-1}$ | 0.97 |
| $J_V$ | Jugular vein-mediated viral transport rate | $\text{d}^{-1}$ | 2.32 |

**Figure 3.**
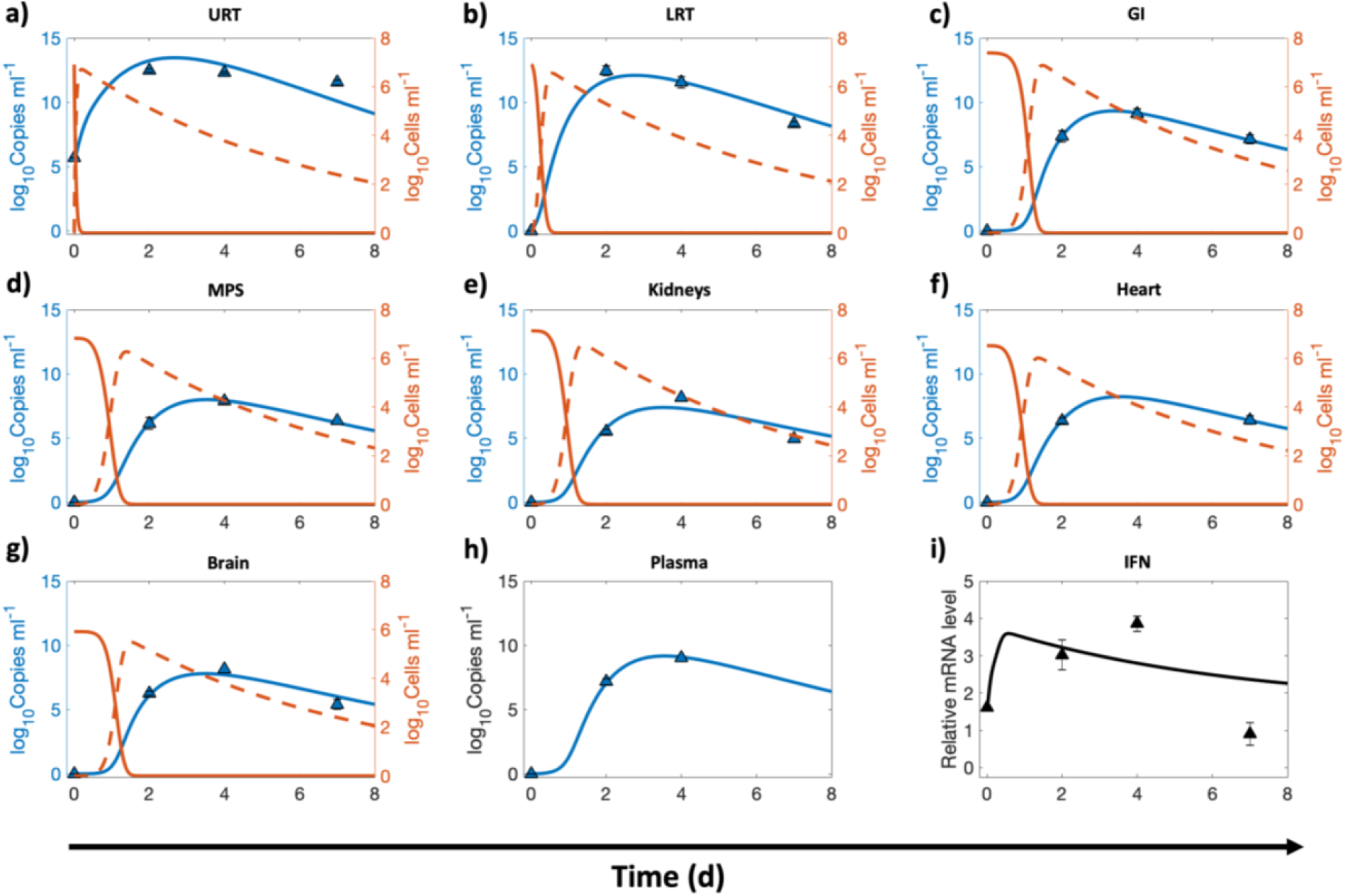
Whole-body reduced model calibration with *in vivo* data. Nonlinear least squares regression of the whole-body reduced model using published *in vivo* data for hamsters. The compartments under consideration are **a**) upper respiratory tract (URT), **b**) lower respiratory tract (LRT), **c**) gastrointestinal tract (GI), **d**) MPS, **e**) kidneys, **f**) heart, **g**) brain, and **h**) plasma. Orange solid and orange dashed lines indicate the population of healthy and infected ACE2-expressing target cells in a given compartment, respectively; blue solid line represents the viral load in the corresponding compartment. Left y-axis represents viral load and right y-axis represents cell populations in **a** through **g. i**) Logarithm base 10 of IFN mRNA copies per *γ*-actin multiplied by 10^6^ is shown. The triangular markers represent experimental data presented as mean ± SD (n = 5 animals per time point). Pearson correlation coefficient between observed and fitted values is *R* > 0.97 (Figure S1a).

The model solution (**Figure 3**) shows the kinetics of ACE2-expressing target cells (solid orange lines) and their infected counterparts (dashed orange lines) in every compartment. These infected cells can produce new virions that will in turn infect other healthy target cells. Because we are using a target-cell limited modeling assumption^9, 18^, the healthy target cells that become infected by the virus are not replaced by new healthy cells, and as seen in **Figure 3**, the target cells were observed to deplete within 48 h post infection. The viral load kinetics (blue curve) is primarily governed by the interplay of new virion production, distribution of the virions between compartments, viral elimination by alveolar and MPS macrophages, hepatobiliary excretion of viruses from the body, cytopathic death of infected cells, and suppression of viral production due to IFN produced by infected cells^9^, which is shown in **Figure 3i**. As the infected cell population tapers, the IFN concentration will also decrease to the pre-infection baseline value. In our model, infected cells of the respiratory tract are the source of IFN following infection, the lack of which has been found to be the underlying cause of life-threatening COVID-19 due to uncontrolled viral replication in the absence of IFN regulation^31-33^.

Of note, the plasma compartment (**Figure 3h**) of the model does not contain any target cell population and thus its viral load kinetics is only governed by the influx and outflux of viruses from various compartments. However, in the full model, the neutralization of viruses by antibodies will also be considered in the plasma compartment, as discussed in the next section. Plasma flow is the key mechanism of viral transport and systemic spread of infection in the body^34^, but due to lack of established mechanistic underpinnings of these processes, we instead use phenomenological rate constants to characterize viral transport. Based on the estimated characteristic times (1-24 h) of the vascular transport processes (shown in **Figure 1** and presented as rates in **Table 2**), it can be inferred that viral transport is permeability-limited and not perfusion-limited, i.e., capillary permeability and vascular surface area govern the rate of extravasation of virions from blood vessels into tissue interstitium to reach the target cells, and thus viral transport is not exclusively governed by the plasma flow rates into the organs. This is consistent with the *in vivo* behavior of nanomaterials of comparable size^35-40^, and is in contrast to the perfusion rate-limited kinetics of smaller lipophilic molecules. The variability in characteristic times of vascular transport can be explained by differences in the permeability of capillary endothelium due to differences in pore sizes of endothelial fenestrae^41^. For instance, the blood brain barrier seems to resist transport of virus to the brain, thereby leading to an estimated characteristic time of influx of 1 day, which is ∼twenty-times longer than the estimated characteristic time of influx to the MPS (1.25 h) that contains large sinusoidal pores in its microvasculature. Of note, the non-vascular transport processes have relatively longer characteristic times that can be attributed to resistance to transport offered by mucus or degradation caused by pH, among other factors.

#### Calibrating parameters of the full model

Once the parameters discussed in the previous section were estimated, they were then used in the full model to calibrate the remaining parameters (see **Table 3**) relevant to the innate and adaptive immune system using published clinical data (*n* = 4 untreated patients)^30^. Due to the uncertainty associated with the duration between day of infection and onset of symptoms (referred to as incubation period), a shifting parameter *τ* was included in the calibration routine. Numerically, the time points corresponding to the data were shifted *τ* units of time. As shown in **Figure 4**, the model correctly represents the initial conditions of the variables and predicts an incubation period *τ* of ∼6 days (indicated by red arrow in **Figure 4a**), which is comparable to published literature^42^. Also, assuming nasal route as the route of infection, the numerical solution for URT at time *t* = 0 suggests exposure to a viral load of ∼10^7^ copies/mL. The clinical data shows the viral load kinetics in the upper (**Figure 4a**) and lower respiratory tract (**Figure 4b**), the IFN kinetics (**Figure 4i**), the effector CD8^+^ (CD8^*^) and activated CD4^+^ (CD4^*^) cell population kinetics (**Figure 4k**), and the total neutralizing antibody kinetics (**Figure 4l**). As shown in **Figure 4**, the full model solution fits the data well (Pearson correlation coefficient *R* > 0.98, *p* < 0.0001, **Figure S1b**), and was able to predict the kinetics of viral load in the remaining compartments by using the viral dynamics and transport parameters estimated from the *in vivo* data (through calibration of the reduced model). The model predicts that the viral load in extrapulmonary organs and plasma persists for ∼17-20 days post onset of symptoms, consistent with published studies^43^, and is thus comparable to the duration of viral detection in URT and LRT. Therefore, it can be also inferred that nasopharyngeal swabs can safely provide an indication of the infection status of the patients.

**Table 3.** List of model parameters estimated from fitting the full model to clinical data.

| Notation | Definition | Units | Value |
| --- | --- | --- | --- |
| **Innate immunity parameters** |  |  |  |
| $P_{\text{IFN}}$ | IFN production rate from infected cells | $\text{pg} \cdot \text{cell}^{-1} \cdot \text{d}^{-1}$ | 5.37 |
| $\epsilon$ | IFN effectiveness | $\text{mL} \cdot \text{pg}^{-1}$ | 4.57 |
| **Viral dynamics parameters** |  |  |  |
| $v_U^0$ | Initial concentration of viral load in the URT | $\text{virus} \cdot \text{mL}^{-1}$ | $10^{6.9}$ |
| $\tau$ | Viral incubation time | d | 5.77 |
| **Adaptive immunity parameters** |  |  |  |
| $T_{\text{APC}}$ | Transition rate of naïve APCs into APC* | $\text{mL} \cdot \text{virus}^{-1} \cdot \text{d}^{-1}$ | 65.2 |
| $T_{\text{T-cells}}$ | Transition rate of naïve CD8 into CD8* or CD4 into CD4* | $\text{mL} \cdot \text{cell}^{-1} \cdot \text{d}^{-1}$ | 0.0061 |
| $T_{\text{BC}}$ | Transition rate of naïve B cells into B* | $\text{mL} \cdot \text{cell}^{-1} \cdot \text{d}^{-1}$ | 0.029 |
| $T_{\text{BC}}^{\text{S}}$ | Transition rate of B* into short-lived plasma cells | $\text{mL} \cdot \text{cell}^{-1} \cdot \text{d}^{-1}$ | 564.8 |
| $T_{\text{BC}}^{\text{L}}$ | Transition rate of B* into long-lived plasma cells | $\text{mL} \cdot \text{cell}^{-1} \cdot \text{d}^{-1}$ | 0.36 |
| $G_{\text{P}}^{\text{S}}$ | Production rate of antibodies from short-lived plasma cells. | $\text{antibody} \cdot \text{cell}^{-1} \cdot \text{d}^{-1}$ | 0.59 |
| $G_{\text{P}}^{\text{L}}$ | Production rate of antibodies from long-lived plasma cells. | $\text{antibody} \cdot \text{cell}^{-1} \cdot \text{d}^{-1}$ | 1.34 |
| $\lambda_{\text{Ab}}$ | Antibody loss rate due to viruses | $\text{mL} \cdot \text{virus}^{-1} \cdot \text{d}^{-1}$ | 0.0013 |
| $D_{\text{Ab}}$ | Elimination rate of viruses due to antibodies | $\text{mL} \cdot \text{antibody}^{-1} \cdot \text{d}^{-1}$ | 0.0085 |
| $D_{\text{CD8}}$ | Death rate of infected cells due to CD8* | $\text{mL} \cdot \text{cell}^{-1} \cdot \text{d}^{-1}$ | 0.0037 |

**Figure 4.**
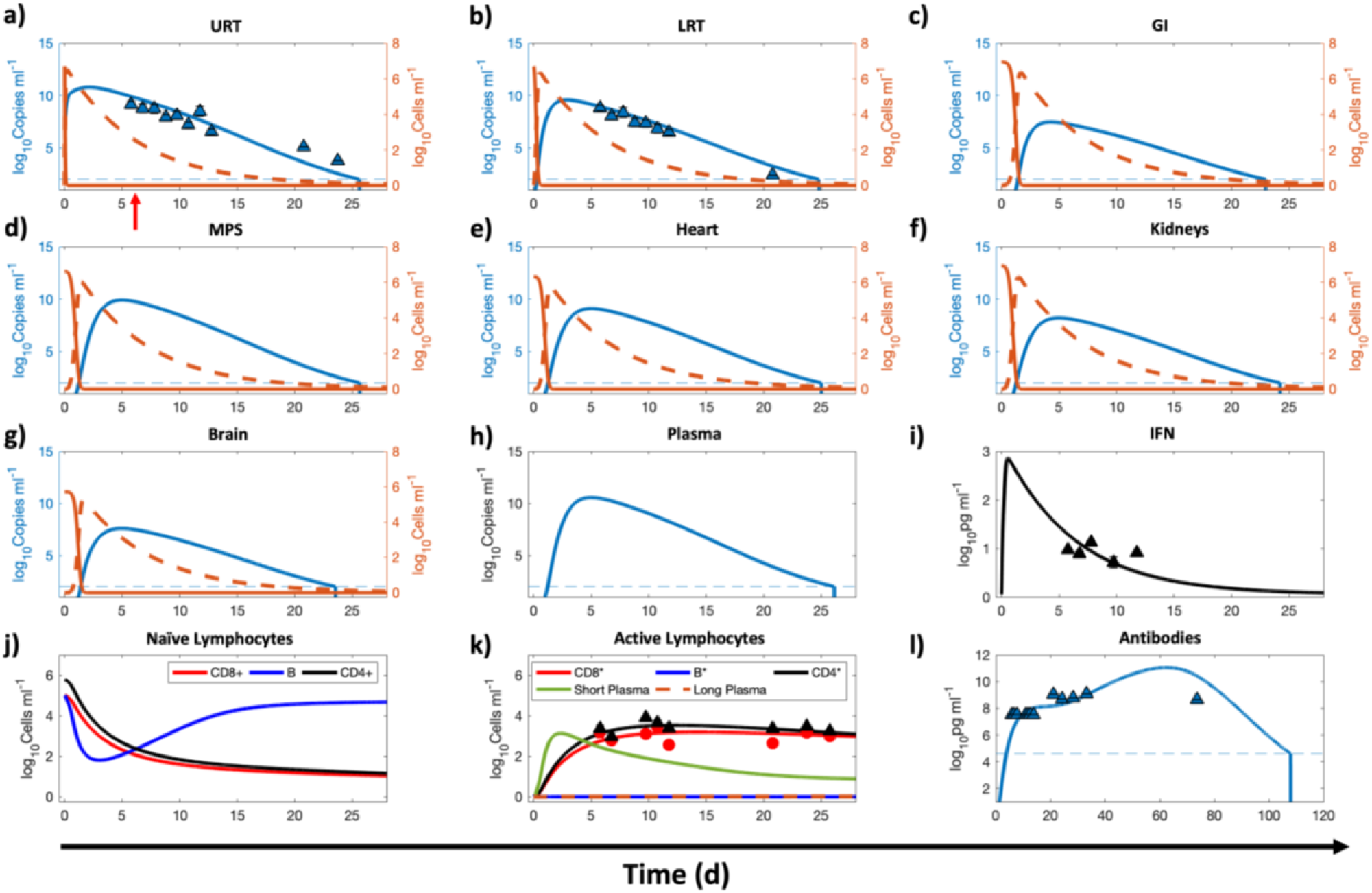
Whole-body full model calibration with clinical data. Numerical results for the full model, corresponding to the second step given in the modeling workflow in Figure 2. The compartments under consideration are **a**) URT, **b**) LRT, **c**) GI, **d**) MPS, **e**) kidneys, **f**) heart, **g**) brain, **h**) blood, and **j), k), l)** lymphatic compartment. Orange solid and orange dashed lines indicate the population of healthy and infected ACE2 expressing target cells in a given compartment, respectively, while the blue solid line represents the viral load in the corresponding compartment. Left y-axis represents viral load and right y-axis represents cell populations in panels **a** through **g. i**) Concentration kinetics of IFN. **j)** and **k)** Kinetics of adaptive immune system cells in the lymphatic compartment. **j)** Naïve/immature CD4^+^, CD8^+^, and B cells. **k)** Cells in their activated/effector state along with antibody producing plasma cells. **l**) Concentration kinetics of total antibody (IgG, IgM, IgA). Red arrow in **a**) indicates time of onset of symptoms. A lower bound at 10^2^ copies ml^-1^ and 10^4.6^ pg ml^-1^ is imposed to represent the detectable limit of viral load and antibodies, respectively; dashed blue line indicates the limit of detection. Once the viral load or antibodies go below the detection limit, a vertical line is used to indicate the time of occurrence of this event. The triangular markers represent clinical data presented as mean ± SD (n = 4 patients). Pearson correlation coefficient between observed and fitted values is *R* > 0.98 (see Figure S1b).

The model also shows the kinetics of naïve lymphocytes (**Figure 4j**) and antibody producing plasma cells (**Figure 4k**) in the lymphatic compartment, which is represented as a common compartment for the entire body. Importantly, in close agreement with published literature^44, 45^, the model predicts that the systemic concentration of antibodies persists above the detectable limit for >100 days post onset of symptoms, following which it may no longer be detectable (**Figure 4l**). This finding suggests the lack of indefinite antibody protection against reinfection^46^, and highlights the need for vaccine boosters to achieve long-lasting immunity^47^. We note that the data used in the above calibrations was obtained under conditions where neither the animals nor the patients were given any pharmacological treatment. Hence, the data are appropriate to calibrate the effects of the immune components and other physiological processes in distributing and eliminating the viral load.

While URT and LRT are the preferred sites to detect the presence of SARS-CoV-2, it is important to note, and as is evident from the model predictions, that the viral load in non-pulmonary organs can attain comparable levels, and can thus explain the non-respiratory symptoms observed in some COVID-19 patients^28, 48, 49^. Following transport of the virus from respiratory tract to blood or via gastrointestinal tract to blood, organs that have a significant population of ACE2 expressing cells may become infected by the virus, leading to the extrapulmonary manifestations of COVID-19 that may include symptoms such as diarrhea and impaired renal-, hepatic-, cardiovascular-, or neurological-functions.

### Individualized effects of immune components on viral kinetics

To investigate the effects of the cellular and humoral arms of innate and adaptive immunity on viral load kinetics in the body, we individually switched off these components and simulated the whole-body viral kinetics for up to the time when viral load fell below the detectable limit^18^ of 10^2^ copies ml^-1^. This numerical experiment is meant to mimic the effect of compromised immunity due to an underlying condition in a virtual patient undergoing no antiviral treatment. As seen in the viral kinetics in **Figure 5**, when one or all of the immune components were shut down, the viral concentration in all the compartments was higher than the baseline (dashed dark red line). Further, it can be inferred that IFN is the primary mechanism of controlling viral load in the URT (**Figure 5a**) and GI (**Figure 5c**), while macrophages (alveolar macrophages, Kupffer cells, and splenic macrophages) play a predominant role in limiting infection in the LRT, MPS, plasma, and other organs, followed by IFNs (**Figures 5b, 5d-5h**). Findings in the literature support the above observations as follows. Lack of IFNs can lead to excessive viral production^33^ and cause life-threating COVID-19 in patients deficient in functional IFNs due to, for example, the occurrence of loss-of-function mutations in genes governing IFN-mediated immunity^32^ or auto-antibodies against IFNs^31^. Further, FABP4+ alveolar macrophages were observed to be largely absent in patients with severe COVID-19, but were a predominant macrophage in patients with mild disease^50^, indicating the major role of FABP4+ alveolar macrophages in controlling infection as also shown previously for patients with chronic obstructive pulmonary disease (COPD)^51^. We assume a constant supply of macrophages in the lungs and MPS in our model, but a deleterious effect of infection on these immune cells cannot be ruled out, and further experimental evidence is necessary to model the cell population kinetics appropriately^52^. Further, the model reveals that the effect of adaptive immunity (antibodies and CD8^*^ cells) is not significant in controlling infection, but it does not necessarily rule out the therapeutic potential of exogenously administered antibodies or novel cell-based therapies (e.g. T cell therapy).

**Figure 5.**
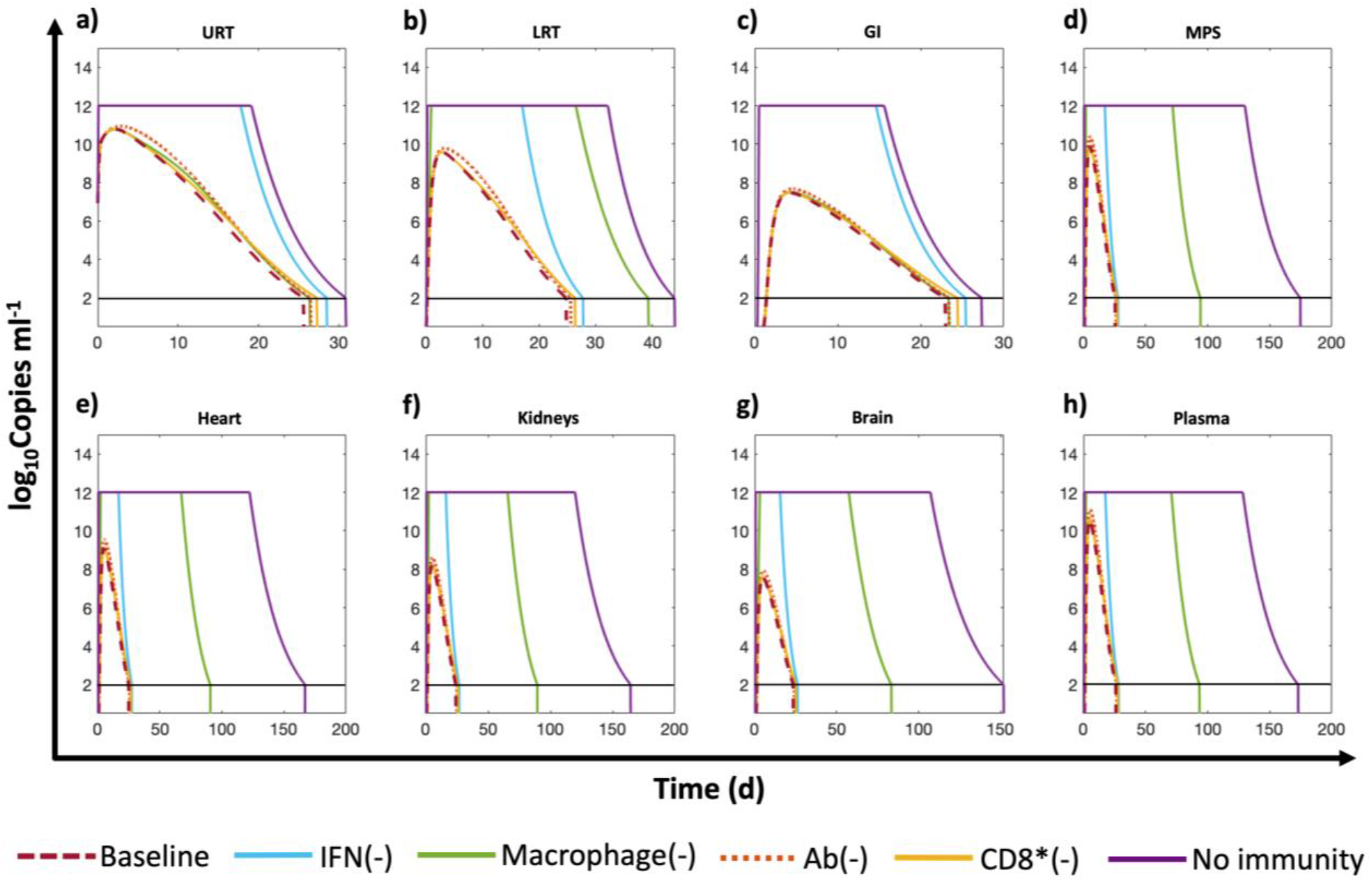
Effects of different components of the immune system. Comparison between different scenarios where a particular mechanism of immunity has been turned off. The viral kinetics in a given compartment is presented on the y-axis. The compartments under consideration are **a**) URT, **b)**LRT, **c**) GI, **d**) MPS, **e**) heart, **f**) kidneys, **g**) brain, and **h**) plasma. Six scenarios are considered. **1)**Baseline: All immune mechanisms are functional. **2)** IFN(-): the innate immune effects of IFN are deactivated. **3)** Macrophage(-): alveolar and MPS macrophages are eliminated. **4)** Ab(-): no antibodies are produced. **5)** CD8^*^(-): cytotoxic effects of CD8^*^ are suppressed. **6)** No immunity: all immunity mechanisms are absent. Flat line at 10^12^ copies ml^-1^ represents the upper bound imposed on viral load to showcase only clinically relevant results. A lower bound at 10^2^ copies ml^-^ 1 is imposed to represent the detectable limit of viral load in the body, indicated by horizontal black line; once the viral load goes below the detection limit, a vertical line is used to indicate the time of occurrence of this event.

All the above scenarios abstractly represent real-world underlying conditions (e.g. cancer, diabetes, autoimmune diseases) in patients that lead to varying degrees of immunosuppression, thus highlighting the importance of an individual’s immune status in regulating viral kinetics. In the absence of immune responses, the only plausible mechanism that brings the viral load down is hepatobiliary excretion of the virus through the MPS, but our results show it takes several weeks before the viral load falls below the detectable value of 10^2^ copies ml^-1^. Note that upon shutting down IFN- and macrophage-mediated immunity or total immunity, the viral load grew beyond clinically observed values in the literature (∼10^12^ copies/ml), therefore we set an upper bound at 10^12^ copies/ml to keep the results clinically meaningful.

### Parametric analysis

To identify the significance of model parameters in affecting viral load kinetics, we performed global sensitivity analysis using the full model (**Figure 6**). The model outputs used to investigate the influence of input model parameter perturbations were area under the curves of viral load kinetics in URT, LRT, and plasma from 0 to 30 days (i.e., 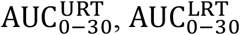, and 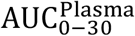, respectively), and time to reduce the viral load to < 10^2^copies mL^-1^ in URT, LRT, and plasma (i.e., 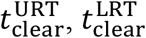, and 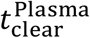, respectively).

**Figure 6.**
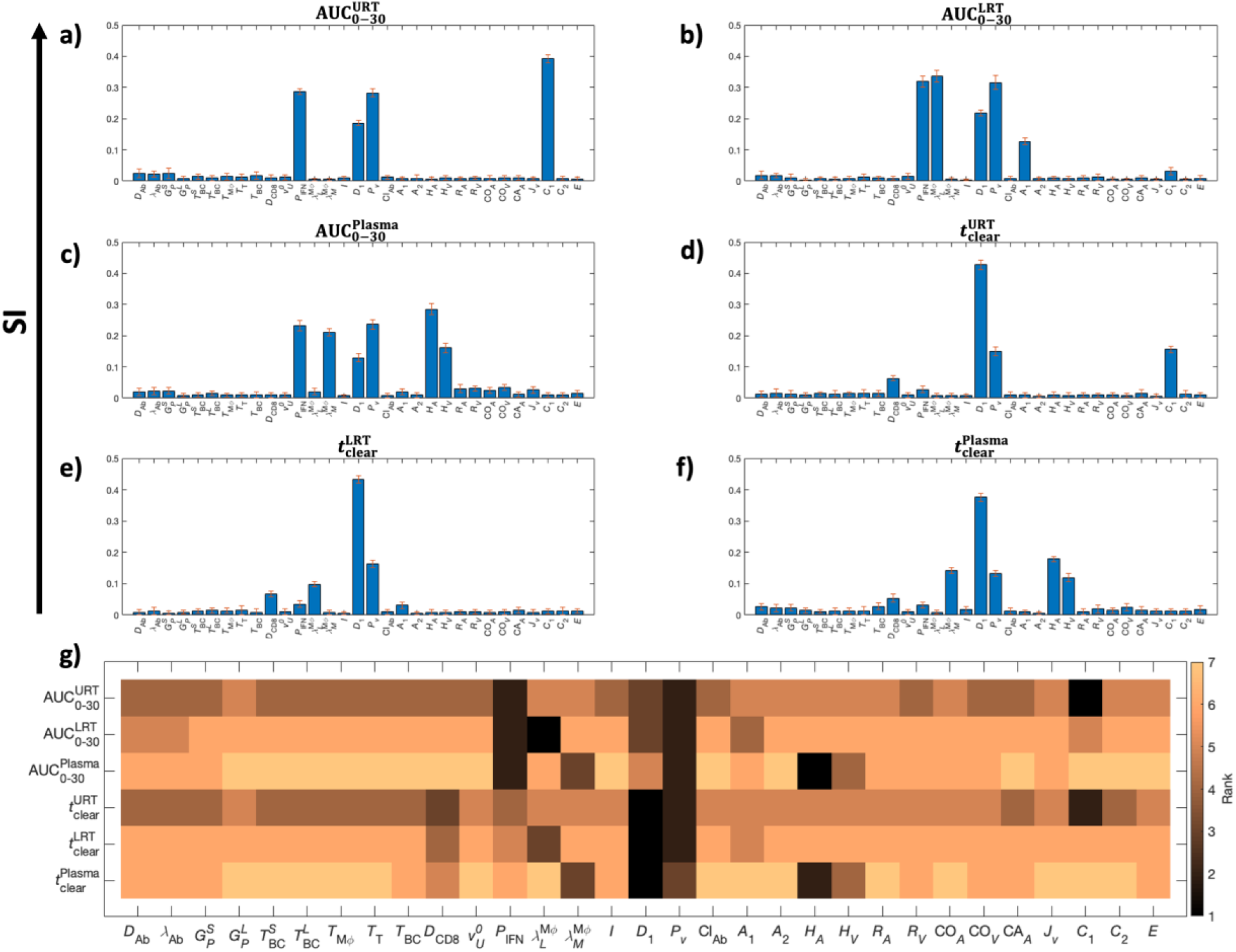
Global sensitivity analysis. All parameters are perturbed simultaneously from their reference value. The ranking of each parameter is quantified as the sensitivity index (SI, y-axis) measured under six criteria. The first three are area under the curves of viral load kinetics 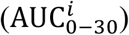 for the first 30 days in the **a)** URT, **b)** LRT, and **c)** plasma compartment. The next three are the total time 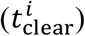 required for the viral load to fall below the detectable concentration of 10^2^ copies ml^-1^ in the **d)** URT, **e)** LRT, and **f)** plasma compartment. Each SI is accompanied with an error bar resulting from the sampling distribution. **g)** Heat map indicating the ranking of the parameters based on the magnitude of the sensitivity index.

As shown in **Figure 6a-f**, multivariate linear regression analysis (MLRA)-based sensitivity indices provide insight into the relative importance of model parameters in governing viral load kinetics. The ranking obtained from MLRA (**Figure 6g**) suggests that the total viral load 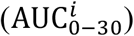 in URT, LRT, and plasma (indicator of systemic behavior) is strongly dependent on viral production rate (*P*_*v*_), IFN production rate (*P*_IFN_), and cytopathic death rate of infected cells (*D*_1_). Cytopathic death of infected cells is a property of the system that may be hard to manipulate pharmacologically, therefore IFN concentration and viral production rate are conceivably the targets best-suited for pharmacological interventions to constrain the viral load in patients^53^. Treatment over a combination of the two parameters, i.e., IFN and viral production rates, can be more efficient than monotherapy in suppressing viral load, as also suggested by clinical studies^54, 55^. While cytopathic death rate and viral production rate play major roles in governing the clearance time of viruses 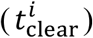, IFN production rate is relatively less significant. Further, transport processes play a significant role in governing both total viral load and time to clear the load. Specifically, the transport of virus from URT to LRT (*C*_1_) affects viral load in URT, and blood to MPS via the hepatic artery (*H*_*A*_) and MPS to blood via the hepatic vein (*H*_*V*_) govern the clearance time from plasma, which should also affect the viral clearance of other organs connected to plasma.

Also, the elimination rate of viruses by alveolar macrophages 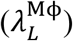 and MPS macrophages 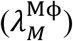 are important parameters in governing the kinetics of viral load in LRT and plasma, respectively. Finally, the total viral load in LRT is also affected by pulmonary absorption (*A*_1_), i.e. by the rate of transport of viruses from LRT to plasma.

### Clinical application of model

Up to this point, we have demonstrated that the model was able to reproduce viral kinetics that were consistent with experimentally measured values and produce observations consistent with published literature. We next sought to examine whether this tool can make predictions that might allow clinicians to design an effective therapy for patients, helping to optimize and personalize their treatment regimen. As a numerical experiment, we tested three treatment scenarios in controlling infection: a hypothetical antiviral agent, interferon therapy, and antiviral agent-interferon combination therapy. Further, the effects of the timing of therapy initiation (*t*^*T*^) were also tested, i.e., starting therapy on the day of onset of symptoms (*t*^*T*^ = *t*^onset^), and starting therapy 5 days post onset of symptoms (*t*^*T*^ = *t*^onset^ + 5). To quantify treatment effectiveness, we compared the viral load kinetics in three compartments, namely, URT, LRT, and plasma in simulations spanning a period of four weeks.

#### Antiviral therapy

We simulated therapy with a *hypothetical antiviral agent that has the same mechanism of action as Remdesivir*^56^, i.e. interference with RNA-dependent RNA polymerase. For this, 200 mg loading dose (≡*i*initial plasma concentration *C*_0_ = 5 *μ*M, assuming a molecular weight of 800 g/mol and a volume of distribution of 50 L), followed by 100 mg daily maintenance doses for 9 additional days, via intravenous injection were simulated to mimic the pharmacological intervention. Treatment was started at one of the two time points previously mentioned. As a simplification, assuming one-compartment pharmacokinetics for the hypothetical drug with elimination rate constant *k*_C1_ = 1 *d*^−1^, we use the plasma concentration kinetics as a surrogate for tissue concentration kinetics of the drug. The plasma concentration *α*(*t*) is thus given by,

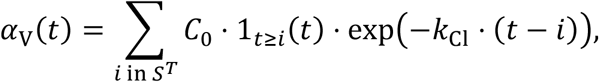

where

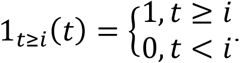

Hence, the injection times are *i* = *t*^*T*^, *t*^*T*^ + 1, …, *t*^*T*^ + 9, and we define this set of times as *S*^*T*^. The antiviral agent acts by inhibiting the production rate *P*_*v*_ of virus from infected cells in the body, such that 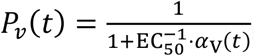, where EC_50_ is the half maximal effective concentration of the drug and is chosen to be 0.77 *μ*M. Plasma concentration kinetics *α*_V_(*t*) of the drug is shown in the inset of **Figure 7a** for the scenario when the treatment was initiated on the day of onset of symptoms.

**Figure 7.**
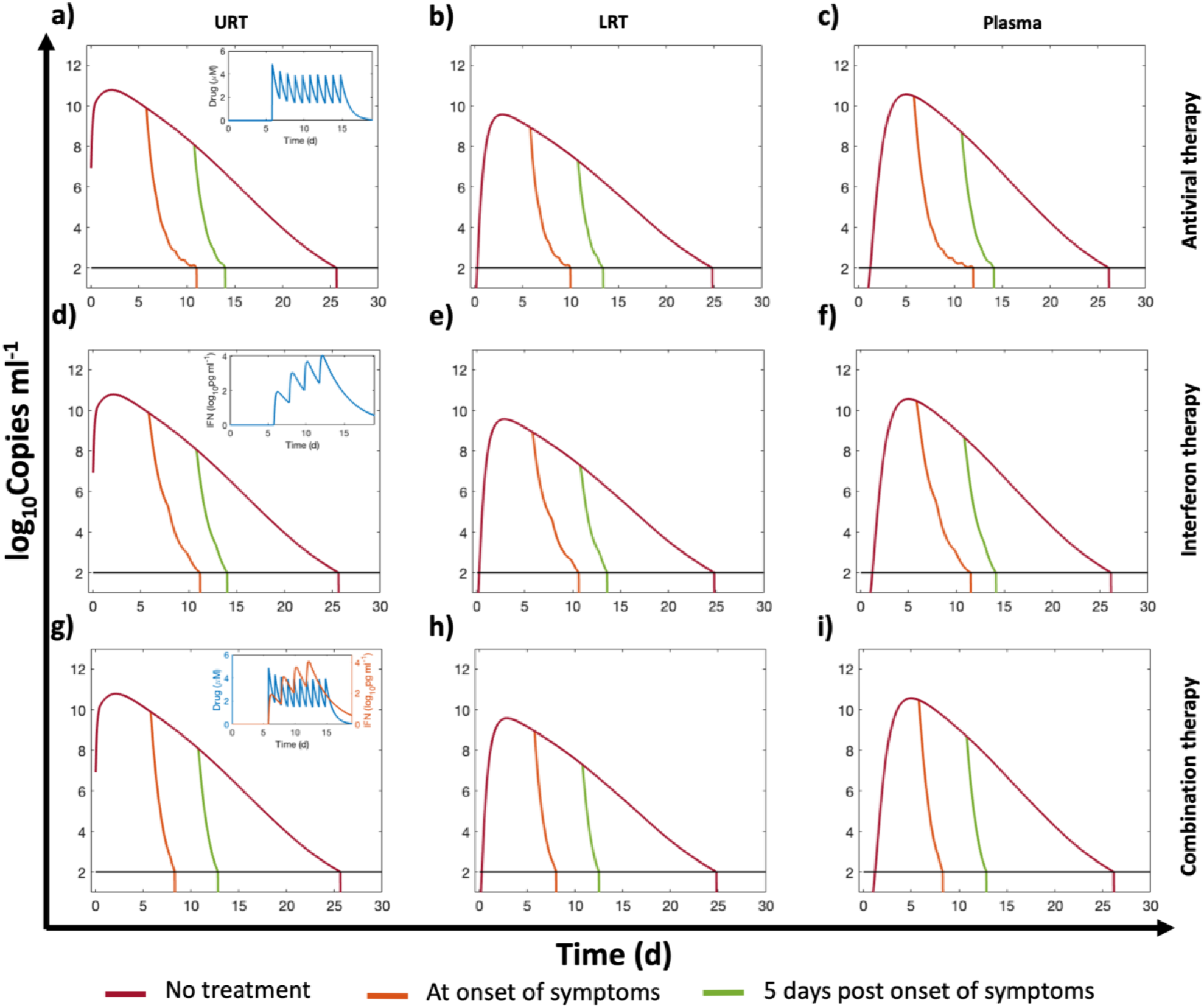
Simulated pharmacological interventions. Viral load kinetics in URT, LRT, and plasma following **a-c)** treatment with a hypothetical antiviral agent, **d-f)** treatment with interferon beta-1a, **g-i)** and combination of the antiviral and interferon beta-1a, starting on the day of onset of symptoms (orange line) and five days post onset of symptoms (green), compared against no treatment (burgundy). Insets in **a**,**d**,**g**) show plasma concentration kinetics of the pharmacological agent/s for the treatment regimen that begins on the day of onset of symptoms. A lower bound at 10^2^ copies ml^-1^ is imposed to represent the detectable limit of viral load in the body (black horizontal line); once the viral load goes below the detection limit, a vertical line is used to indicate the time of occurrence of this event.

As shown in **Figure 7a-c**, under the considered therapeutic regimen, the antiviral therapy shows a significant impact on viral load reduction compared to the no treatment scenario, irrespective of the timing of initiation of therapy. However, an early initiation of antiviral therapy led to a lesser total viral load and a faster reduction of the load, which is consistent with results of other studies^18^.

#### Interferon therapy

We simulated treatment with interferon beta-1a, with and without the hypothetical antiviral agent (discussed above). For this, four subcutaneous injections (Dose = 44 µg each) of interferon beta-1a were administered every other day starting at one of the two time points previously mentioned. The plasma concentration kinetics of interferon beta-1a *α*_I_(*t*) is obtained by solving the following equation:

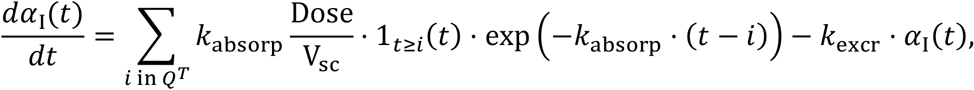

where

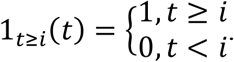

Hence, the injection times are *i* = *t*^*T*^, *t*^*T*^ + 2, …, *t*^*T*^ + 6, and we define this set of times as *Q*^*T*^. The plasma concentration kinetics *α*_I_(*t*) of interferon beta-1a is shown in the inset of **Figure 7d** for the scenario when the treatment was initiated on the day of onset of symptoms. Here, the parameters *k*_absorp_, V_sc_, and *k*_excr_, which represent the absorption of interferon beta-1a from the site of injection to the blood stream, volume of the injection site, and excretion rate of interferon beta-1a, respectively, were estimated by fitting the above equation to literature derived concentration kinetics data^57^. The estimated parameter values for *k*_absorp_ = 7.04 d^-1^, V_sc_ = 3.43 ml, and *k*_excr_ = 0.074 d^-1^. Of note, while interferon beta-1a acts in the same way as endogenous IFN, i.e. by inhibiting the production rate *P*_*v*_ of virus from infected cells, however we separately model them to accommodate the possibility of unique pharmacokinetic properties of the two agents. Therefore, the first term of viral load kinetics in the ODE for organ *i* (see Methods) now becomes:

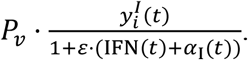

As shown in **Figure 7d-f**, interferon beta-1a therapy significantly reduces the viral load, and the effect is comparable to the hypothetical antiviral agent. Also, an early initiation of therapy leads to a lesser total viral load and a faster reduction in viral load.

#### Combination therapy

Finally, we tested the effect of combination therapy with the hypothetical antiviral agent and interferon beta-1a on viral load kinetics (plasma concentration kinetics in the inset of **Figure 7g**). As shown in **Figure 7g-i**, the combination therapy leads to a faster reduction in viral load compared to the effect of antiviral agent and interferon beta-1a alone, such that the viral load seems to fall below the detection threshold 2-3 days earlier.

## Conclusions

In summary, we have developed a semi-mechanistic mathematical model that predicts the whole-body viral distribution kinetics of SARS-CoV-2 by incorporating cellular-scale viral dynamics, relevant physiological processes of viral transport, innate and adaptive immune response, and viral excretion. The model is well calibrated with published *in vivo* and clinical data and provides insights into the importance of various components of the immune system in controlling infection. Through global sensitivity analysis, we identified the key mechanisms that control infection and can be used as potential therapeutic targets for pharmacological intervention. Finally, we tested the potential of such therapeutic targets by simulating clinically relevant treatment options and identified the importance of the timing of treatment initiation, and the effects of various therapies to suppress infection effectively and immediately. As a limitation of the model, it is important to note that due to lack of clinical data for the viral load kinetics in extrapulmonary compartments, we had to rely on the transport parameters estimated from *in vivo* data, and the model predictions for the extrapulmonary compartments could not be clinically validated. As clinical knowledge of the disease and its mechanisms improves, we will continue to fine-tune the model and integrate it with a physiologically-based pharmacokinetic model to build an in-silico platform that can be used to simulate disease progression and a more complete pharmacokinetics of various test drugs and novel formulations.

## Methods

### 1. Model development

In each organ compartment, we consider the kinetics of three populations. The first is the concentration of healthy cells, denoted by 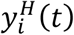, where *i* is an arbitrary compartment. The second is the concentration of infected cells, denoted by 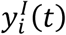, and the third is the concentration of viral particles *v*_*i*_(*t*). The compartments that we consider are *i* = [*U, L, G, M, H, K, B*], where *U* is the upper respiratory tract (URT), *L* is the lower respiratory tract (LRT), *G* is the gastrointestinal tract (GI), *M* is the mononuclear phagocytic system (MPS), which encompasses the liver and the spleen, *H* is the heart, *K* denotes the kidneys, and *B* is the brain. All organ compartments are connected via the plasma compartment *P*(*t*), in which we only consider the concentration of viral particles in systemic circulation.

We assume that, due to the time scale under consideration, the population of healthy cells does not replenish during the simulation window. Thus, the rate of change of healthy cells is modeled as a decreasing function, which depends on the interaction between healthy cells and the surrounding viral particles that infect them. The conversion rate of healthy cells to infected cells is denoted by the infection rate *I*.

The rate of change of total infected cell population within an organ is a function of three mechanisms. The first is an increase due to freshly infected healthy cells by viral particles, the second is death due to cytopathic effect of viral infection, proportional to the number of infected cells and characterized by the rate constant *D*_1_, and the third is due to the interaction between effector CD8^+^ cells (concentration denoted by CD8^*^(*t*)) and the infected cells. The cytotoxic effect is measured in terms of the death rate constant *D*_CD8_. It is important to note that in the reduced form of the model, infected cell death due to effector CD8^*^ cells is not included. In our model description, all activated immune cells are indicated with an asterisk (e.g., CD8^*^), while inactive or Naïve populations are indicated by standard naming conventions (e.g., CD8^+^).

The concentration of viral particles in each organ compartment is influenced by different factors, some of which are organ-specific. However, in all organ compartments, the rate of change of viral particles is proportional to the number of infected cells with virus production constant *P*_*v*_, but is inversely proportional to the concentration of IFN, denoted IFN(*t*). IFN is part of the innate immune response that acts by suppressing the viral production rate of infected cells, and is controlled by the effectiveness constant *ε*. In all compartments, viral particles may be neutralized due to the aggregation of antibodies on their surface proteins. The antibody concentration is represented by Ab(*t*), and the destruction rate of viral particles due to antibodies is denoted by *D*_Ab_. Of note, neutralization of virus by antibodies is not incorporated in the reduced form of the model. Lastly, the viral load can also be reduced by tissue resident macrophages in the lungs (alveolar macrophages) and MPS (Kupffer cells and splenic macrophages) that engulf the viral particles and destroy them. The rate of removal of viral particles by macrophages is given by 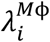.

We introduce the system of ordinary differential equations that governs the concentration kinetics as follows. Mechanisms particular to each compartment are discussed after each set of equations.

Equations for the URT:

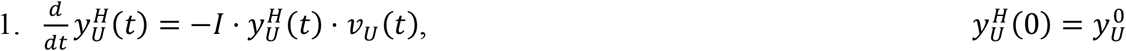

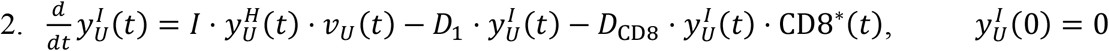

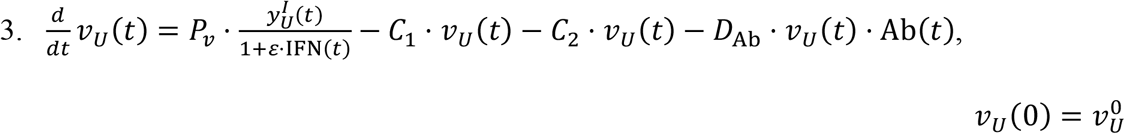

Note that the rate of change of viral particles in the URT depends on two clearance mechanisms. These consist on the migration of viral particles from the URT to the LRT and from the URT to the GI tract. The corresponding transport coefficients are *C*_1_ and *C*_2_, respectively.

Equations for the LRT:

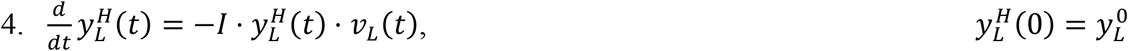

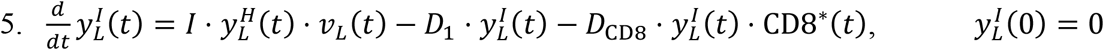

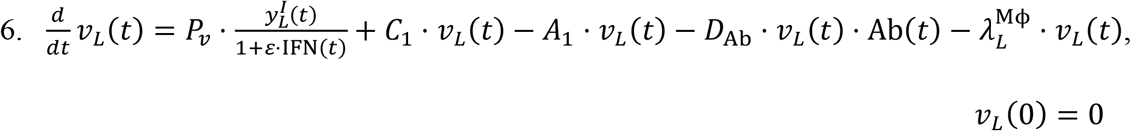

Viral particles in the LRT are received from the URT, but are also lost to the systemic circulation, which is quantified through the pulmonary absorption coefficient *A*_1_.

Equation for IFN:

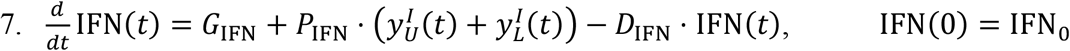

As mentioned earlier, IFN limits the production of viral particles by infected cells. The rate of production of this cytokine is regulated by two effects. One is a zero-th order generation term *G*_IFN_, and the second is proportional to the cumulative population of infected cells in the respiratory tract. The proportionality constant in the second mechanism is *P*_IFN_. Further, IFN can degrade over time at a rate given by *D*_IFN_.

Equations for the GI tract:

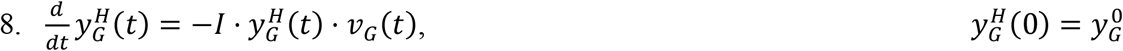

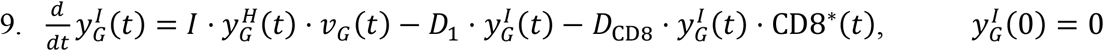

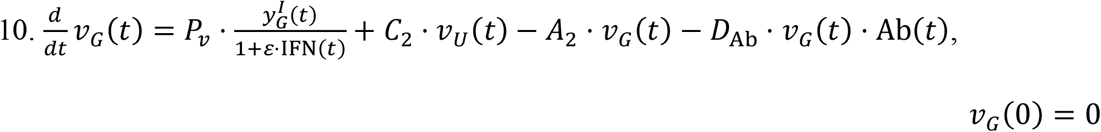

Within the GI tract, in addition to the standard mechanisms, viral particles can be lost to the liver via the hepatic portal vein. This rate is quantified through the intestinal absorption rate *A*_2_. Additionally, as discussed above, viral particles from the URT are transported to the GI tract at a rate *C*_2_.

Equations for the MPS:

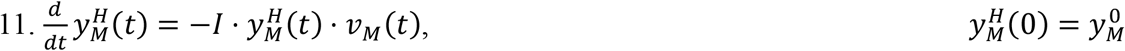

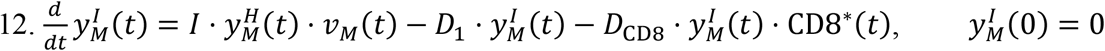

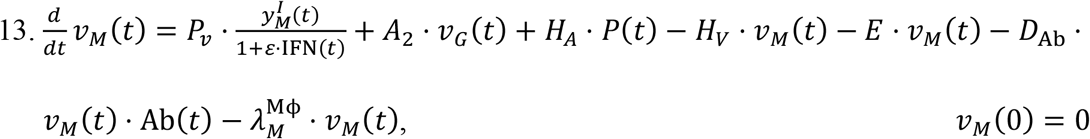

The rate of change of viral particles within the MPS can increase due to the particles collected from the GI tract (*A*_2_) and those incoming from systemic circulation through the hepatic artery (*H*_*A*_). At the same time, particles can leave this compartment through the hepatic vein (*H*_*V*_), or through the hepatobiliary excretion mechanism (*E*).

Equations for the Heart:

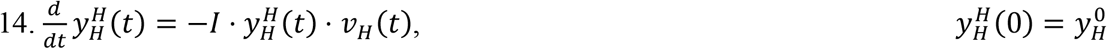

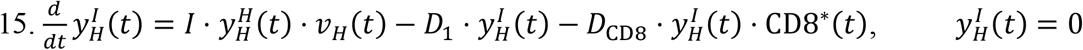

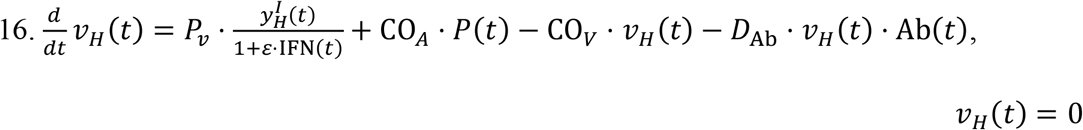

Similar to the MPS, the rate of change of viral particles in the heart depend on two fluxes. One is the incoming viral load from systemic circulation through the coronary arteries (CO_*A*_), and the second is the outgoing viral particles via the coronary veins (CO_*V*_).

Equations for the Kidneys:

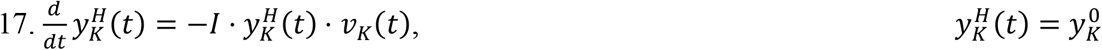

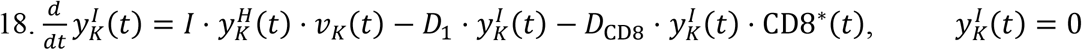

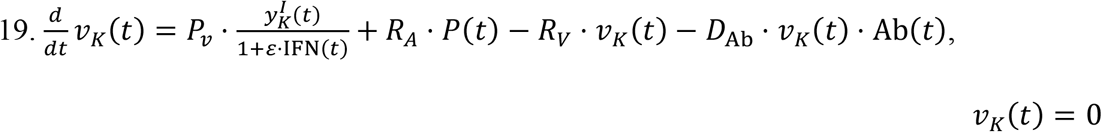

Given that the size of the SARS-CoV-2 virus is ∼100 nm, we assume that renal excretion is not feasible^58^. Therefore, the viral kinetics in kidneys is dependent on the standard mechanisms and is affected by influx via the renal arteries (*R*_*A*_) and outflux via the renal veins (*R*_*V*_).

Equations for the Brain:

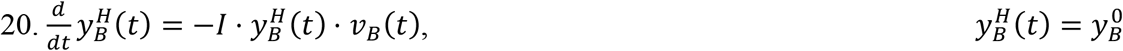

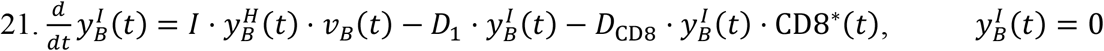

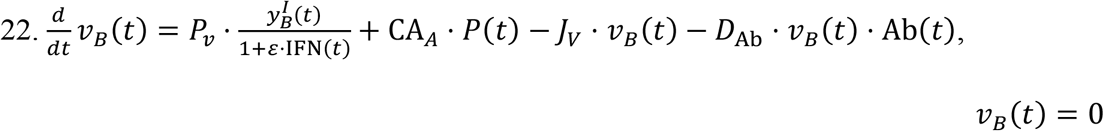

While the blood brain barrier may be deterrent to the establishment of infection in the brain, we have included the brain compartment due to the lack of evidence for the former^59^. Analogous to the heart and kidney compartments, the brain receives viral particles delivered through systemic circulation via the internal carotid arteries (CA_*A*_). Subsequently, viral particles can rejoin the plasma compartment by means of the internal jugular veins (*J*_*V*_).

Equations for Plasma:

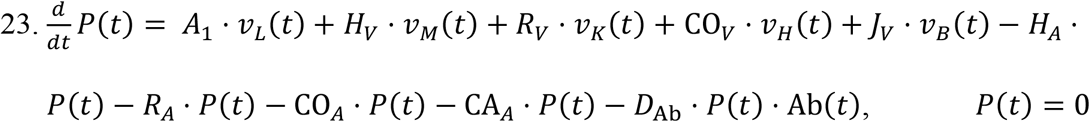

The equation for the plasma compartment incorporates all the outgoing fluxes of viral particles through arteries (*H*_*A*_, *R*_*A*_, CO_*A*_, CA_*A*_) and the incoming fluxes via veins (*H*_*V*_, *R*_*V*_, CO_*V*_, *J*_*V*_). Also, the viral load from LRT in equation (6) gets added to plasma at rate *A*_1_. Lastly, similar to all the previous compartments, viral particles can be neutralized by antibodies at a rate *D*_Ab_.

The model up to this point is the **reduced model**, and the only equation that captures the immune system is equation (7). This equation describes the change of concentration of IFN, which is part of the innate immune system. However, the adaptive immune system should activate to properly mount a full immune response. The equations that follow provide the remaining elements to initiate and maintain a humoral and cell-mediated adaptive immune response to make up the **full model**.

Equations (24)-(30) model the concentration of naïve antigen presenting cells (APCs) APC_*i*_(*t*) in a given compartment *i*. These cells predominantly act as carriers, transporting remnants of viral particles to the lymphatic compartment to raise the alarm in the adaptive arm of the immune system. In order to keep the population of APCs at steady state, a replenishing mechanism is included. It is proportional to the difference between the current concentration APC_*i*_(*t*), and the generic steady state value 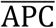, divided by the absolute value of the same difference plus a modulating constant that we set = 1, i.e., 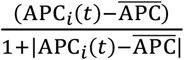. The rationale behind the previous definition is that if the concentration APC_*i*_(*t*) is close to 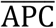 the term is essentially zero, whereas if the difference is large, the denominator is equally large, and the ratio is close to one. This allows a near constant modulation rate that only acts whenever the population of macrophages is far from equilibrium. This reequilibration rate is denoted by *G*_APC_. The APC population is also impacted by the interaction between APCs and invading viral particles. This causes the population of APCs to become activated at a rate *T*_APC_, which is proportional to the product of the concentration of these cells and the viral load.

Equations for URT Naïve APCs:

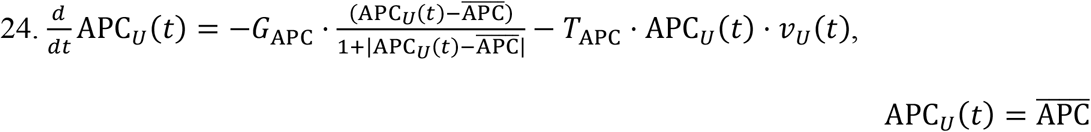

Equations for the LRT Naïve APCs:

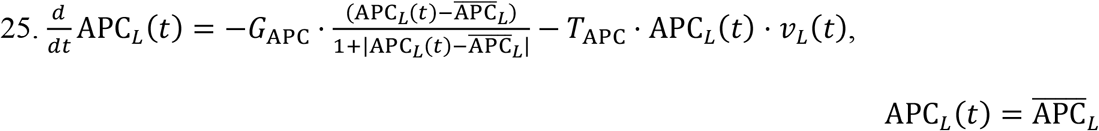

GI tract Naïve APCs:

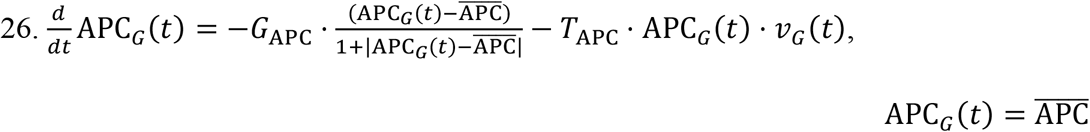

MPS Naïve APCs:

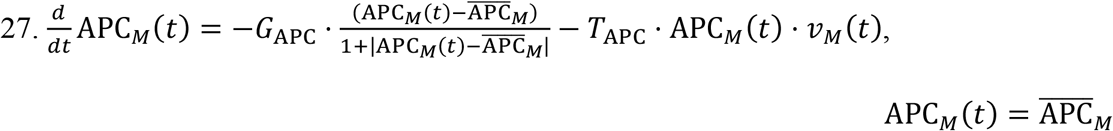

Heart Naïve APCs:

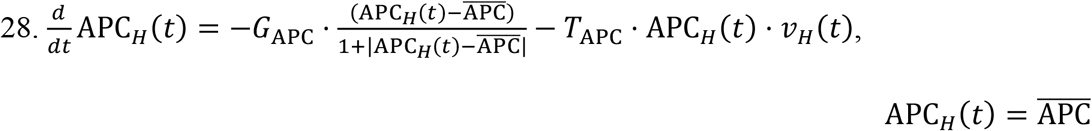

Kidney Naïve APCs:

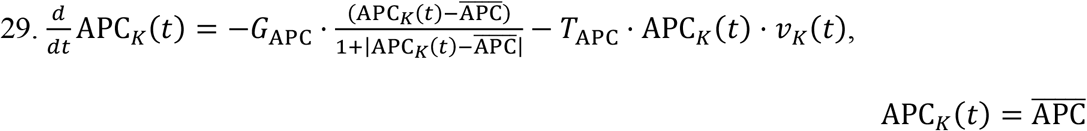

Brain Naïve APCs:

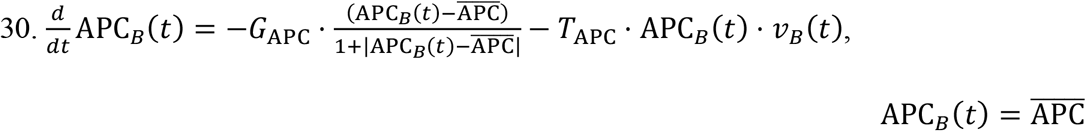

Lymphatic Activated APCs:

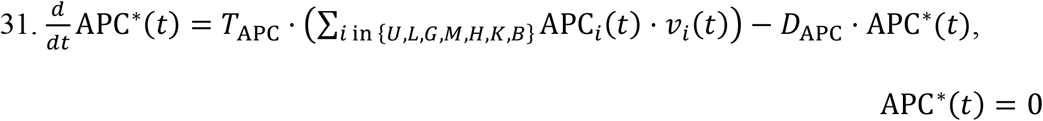

As mentioned earlier, the concentration of activated APCs, denoted by APC^∗^(*t*), increases at a rate proportional to the conversion rate *T*_APC_ of naïve APCs to the activated state. At the same time, activated APCs can become incapacitated or simply eliminated after a certain period of time. These processes are pooled into the APC death rate constant *D*_APC_.

The remaining equations in the model describe either lymphocyte or antibody concentrations. The lymphocytes under consideration are CD8^+^, CD4^+^, and B cells. The concentration of the naïve or inactivated version of these populations is represented by CD8(*t*), CD4(*t*), and BC(*t*), respectively. Similar to the APC case, the steady state concentration for each of these cell types is denoted with a horizontal bar on top of each variable. This value is maintained using a mechanism analogous to the APC case, i.e., through the term 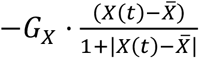, where *X* is the corresponding cell type.

Furthermore, each of these lymphocytes can be promoted to its activated state, which we denote with an asterisk, *X*^∗^. This conversion takes place when activated APCs interact with naïve lymphocytes, presenting them fragments of the ingested viral particles. The transformation rate from the naïve state to the activated one is represented by the constant *T*_*X*_, where *X* indicates the cell type. For the case of CD4^+^ and CD8^+^, we use the common rate *T*_T_. The concentration of activated lymphocytes can fluctuate due to two mechanisms. One is the conversion of naïve cells, which was already discussed. The second is due to death caused by a variety of factors like exhaustion or apoptosis, and is expressed through the death constant *D*_*X*_. B cells, however, follow a different mechanism, which we discuss later.

Naïve CD8:

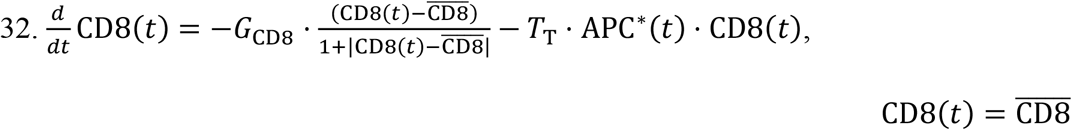

Active CD8^*^:

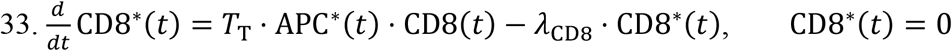

Naïve B cells:

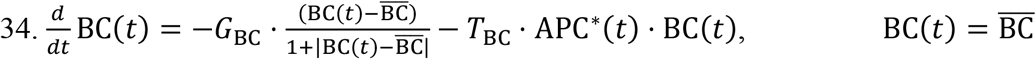

Activated B cells:

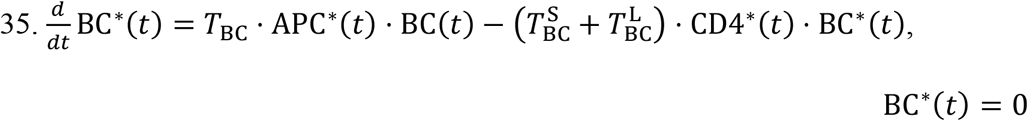

Activated B cells alone cannot neutralize the viral load. They must first transform into plasma cells, which then in turn produce antibodies that can carry out the viral neutralization. The transition from activated B cell to plasma cell is enabled by activated CD4^*^ cells. This transformation can generate two types of plasma cell: short-lived (S) and long-lived (L). The transition rates are represented by the constants 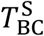 and 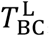, and the corresponding concentrations of plasma cells is denoted by P^S^(*t*) and P^L^(*t*), respectively. Lastly, each type possesses a particular death rate, indicated by the constant 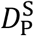 for the short-lived type and 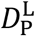 for the long-lived type.

Naïve CD4:

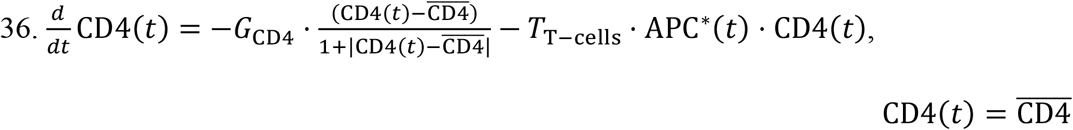

Activated CD4:

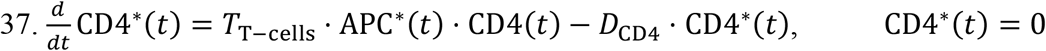

Short-lived plasma cells:

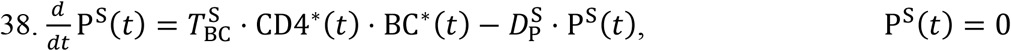

Long-lived plasma cells:

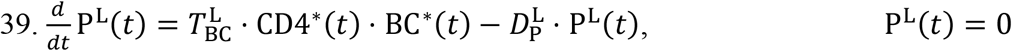

Antibodies:

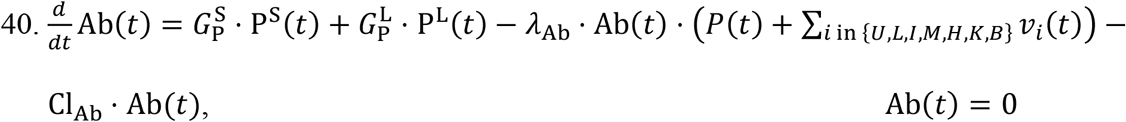

The final equation in the model describes the rate of change of antibody concentration, Ab(*t*). The sole contributors to the production of antibodies are short-lived and long-lived plasma cells. The corresponding production rates are given by 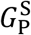 and 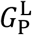, respectively. We consider two mechanisms by which antibodies can be consumed or eliminated. One is the loss of antibodies due to their interaction with viral particles in a given compartment with rate constant *λ*_Ab_. The second is the elimination of antibodies by other factors independent of the viral load, e.g., degradation or clearance from tissues, which occurs at rate constant CL_Ab_. A summary of the parameters used in the model is given in **Tables 1-3**.

### 2. Parametric analysis

Once the full set of parameters for the model has been defined and fixed for the conditions corresponding to **Figure 4**, we proceed to identify the relative effect of each parameter on the viral kinetics. This is done by systematically perturbing one or multiple parameters and comparing the outcomes between the perturbed state and the original state. To quantify the differences between the two states we introduce the following 6 criteria. The first three refer to the area under the curve (AUC) of the viral load curves in the URT, the LRT, and the Plasma compartment for a period of 30 days. Mathematically,

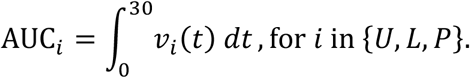

The next three denote the total time required for the viral load in the URT, LRT, and Plasma compartment to reduce to < 10^2^ viral copies per mL, i.e., 2 in log base 10 units. The last comparison criterion is the antibody titer at day 30 in the body, i.e., Ab(*t* = 30). In the next two subsections we compare each of the 6 criteria using global sensitivity analysis (GSA), applied to the 31 parameters.

#### Global sensitivity analysis

Local sensitivity analysis only allows one parameter to change at a time, and thus may not be suitable for analyzing complex biological systems. We performed global sensitivity analysis (GSA^36, 54, 55^; i.e., multiple parameters can be varied simultaneously) to obtain a further understanding of the relationship between parameters and their combined effects on the model outputs of interest. Specifically, in our analysis, all parameters were perturbed at the same time, and we assume a uniform distribution for each parameter. The range of values of each sample was PR ± 99%(PR), where PR is the reference value of a given parameter. To ensure a complete sampling of the full parameter space we used a Latin hypercube sampling scheme. Subsequently, each sample was used to recompute the kinetics, from which we extract the 6 criteria mentioned above. In total, 10 batches of 5000 samples each were computed. Then, the data were subjected to a multivariate linear regression analysis to produce an impact factor, which we label as the sensitivity index (SI). Lastly, the sensitivity indices were ranked by means of a one-way ANOVA and a Tukey’s test. The higher the SI, the more significance a parameter holds. A detailed description of the GSA workflow can be found in^38, 60, 61^. All analyses were performed in MATLAB R2018a.

## Data Availability

The manuscript uses and references data from published literature.

## Author Contributions

PD, ZW conceived the study, conceptualized the idea, and designed the model. PD, JRR developed the model. PD, JRR, KS, MJP, MR collected the data. PD, KS, JDB, VKY, RP, WA, MB, DS, VC, ZW interpreted the data. PD, JRR, JDB, ZW performed the model analysis. PD, JRR, KS, JDB, MJP, MR, RP, WA, DS, VC, ZW wrote or edited the manuscript.

## Acknowledgements

This research has been supported in part by the National Science Foundation Grant DMS-1930583 (VC, ZW), the National Institutes of Health (NIH) Grants 1U01CA196403 (VC, ZW), 1U01CA213759 (VC, ZW), 1R01CA226537 (RP, WA, VC, ZW), 1R01CA222007 (VC, ZW), and U54CA210181 (VC, ZW). The funders had no role in study design, data collection and analysis, decision to publish, or preparation of the manuscript.

## Competing Interests

The authors have declared that no competing interests exist.

## Notes

### Competing Interest Statement

The authors have declared no competing interest.

### Summary of Updates

Author affiliations updated

## References

1. Evans, O., Socio-economic impacts of novel coronavirus: The policy solutions. BizEcons Quarterly 2020, 7, 3–12.

2. Barry, M., Al Amri, M., Memish, Z. A., COVID-19 in the Shadows of MERS-CoV in the Kingdom of Saudi Arabia. Journal of Epidemiology and Global Health 2020, 10 (1), 1–3.

3. Wrapp, D., Wang, N., Corbett, K. S., Goldsmith, J. A., Hsieh, C.-L., Abiona, O., Graham, B. S., McLellan, J. S., Cryo-EM structure of the 2019-nCoV spike in the prefusion conformation. Science 2020, 367 (6483), 1260–1263.

4. Hamming, I., Timens, W., Bulthuis, M., Lely, A., Navis, G., van Goor, H., Tissue distribution of ACE2 protein, the functional receptor for SARS coronavirus. A first step in understanding SARS pathogenesis. The Journal of Pathology: A Journal of the Pathological Society of Great Britain and Ireland 2004, 203 (2), 631–637.

5. To, K. K.-W., Tsang, O. T.-Y., Leung, W.-S., Tam, A. R., Wu, T.-C., Lung, D. C., Yip, C. C.-Y., Cai, J.-P., Chan, J. M.-C., Chik, T. S.-H., Temporal profiles of viral load in posterior oropharyngeal saliva samples and serum antibody responses during infection by SARS-CoV-2: an observational cohort study. The Lancet Infectious Diseases 2020.

6. Wang, Y., Heiland, R., Craig, M., Davis, C. L., Versypt, A. N. F., Jenner, A., Ozik, J., Collier, N., Cockrell, C., Becker, A., Rapid community-driven development of a SARS-CoV-2 tissue simulator. BioRxiv 2020.

7. Zitzmann, C., Kaderali, L., Mathematical analysis of viral replication dynamics and antiviral treatment strategies: from basic models to age-based multi-scale modeling. Frontiers in microbiology 2018, 9, 1546.

8. Hancioglu, B., Swigon, D., Clermont, G., A dynamical model of human immune response to influenza A virus infection. Journal of theoretical biology 2007, 246 (1), 70–86.

9. Baccam, P., Beauchemin, C., Macken, C. A., Hayden, F. G., Perelson, A. S., Kinetics of influenza A virus infection in humans. Journal of virology 2006, 80 (15), 7590–7599.

10. Lee, H. Y., Topham, D. J., Park, S. Y., Hollenbaugh, J., Treanor, J., Mosmann, T. R., Jin, X., Ward, B. M., Miao, H., Holden-Wiltse, J., Simulation and prediction of the adaptive immune response to influenza A virus infection. Journal of virology 2009, 83 (14), 7151–7165.

11. Layden, T. J., Layden, J. E., Ribeiro, R. M., Perelson, A. S., Mathematical modeling of viral kinetics: a tool to understand and optimize therapy. Clinics in liver disease 2003, 7 (1), 163–178.

12. Koelle, K., Farrell, A. P., Brooke, C. B., Ke, R., Within-host infectious disease models accommodating cellular coinfection, with an application to influenza. Virus evolution 2019, 5 (2), vez018.

13. Kucharski, A. J., Russell, T. W., Diamond, C., Liu, Y., Edmunds, J., Funk, S., Eggo, R. M., Sun, F., Jit, M., Munday, J. D., Early dynamics of transmission and control of COVID-19: a mathematical modelling study. The lancet infectious diseases 2020.

14. Berhe, B., Legese, H., Degefa, H., Adhanom, G., Gebrewahd, A., Mardu, F., Tesfay, K., Welay, M., Negash, H., Global epidemiology, pathogenesis, immune response, diagnosis, treatment, economic and psychological impact, challenges, and future prevention of COVID-19: A scoping review. medRxiv 2020.

15. Eletreby, R., Zhuang, Y., Carley, K. M., Yağan, O., Poor, H. V., The effects of evolutionary adaptations on spreading processes in complex networks. Proceedings of the National Academy of Sciences 2020, 117 (11), 5664–5670.

16. Zhao, S., Lin, Q., Ran, J., Musa, S. S., Yang, G., Wang, W., Lou, Y., Gao, D., Yang, L., He, D., Preliminary estimation of the basic reproduction number of novel coronavirus (2019-nCoV) in China, from 2019 to 2020: A data-driven analysis in the early phase of the outbreak. International journal of infectious diseases 2020, 92, 214–217.

17. Wu, J. T., Leung, K., Leung, G. M., Nowcasting and forecasting the potential domestic and international spread of the 2019-nCoV outbreak originating in Wuhan, China: a modelling study. The Lancet 2020, 395 (10225), 689–697.

18. Goyal, A., Cardozo-Ojeda, E. F., Schiffer, J. T., Potency and timing of antiviral therapy as determinants of duration of SARS CoV-2 shedding and intensity of inflammatory response. medRxiv 2020.

19. Perelson, A. S., Neumann, A. U., Markowitz, M., Leonard, J. M., Ho, D. D., HIV-1 dynamics in vivo: virion clearance rate, infected cell life-span, and viral generation time. Science 1996, 271 (5255), 1582–1586.

20. Sahoo, S., Hari, K., Jhunjhunwala, S., Jolly, M. K., Mechanistic modeling of the SARS-CoV-2 and immune system interplay unravels design principles for diverse clinicopathological outcomes. bioRxiv 2020.

21. Ke, R., Zitzmann, C., Ribeiro, R. M., Perelson, A. S., Kinetics of SARS-CoV-2 infection in the human upper and lower respiratory tracts and their relationship with infectiousness. medRxiv 2020.

22. Puelles, V. G., Lütgehetmann, M., Lindenmeyer, M. T., Sperhake, J. P., Wong, M. N., Allweiss, L., Chilla, S., Heinemann, A., Wanner, N., Liu, S., Braun, F., Lu, S., Pfefferle, S., Schröder, A. S., Edler, C., Gross, O., Glatzel, M., Wichmann, D., Wiech, T., Kluge, S., Pueschel, K., Aepfelbacher, M., Huber, T. B., Multiorgan and Renal Tropism of SARS-CoV-2. N Engl J Med 2020, 383 (6), 590–592.

23. Zou, X., Chen, K., Zou, J., Han, P., Hao, J., Han, Z., Single-cell RNA-seq data analysis on the receptor ACE2 expression reveals the potential risk of different human organs vulnerable to 2019-nCoV infection. Frontiers of medicine 2020, 1–8.

24. Ziegler, C. G., Allon, S. J., Nyquist, S. K., Mbano, I. M., Miao, V. N., Tzouanas, C. N., Cao, Y., Yousif, A. S., Bals, J., Hauser, B. M., SARS-CoV-2 receptor ACE2 is an interferon-stimulated gene in human airway epithelial cells and is detected in specific cell subsets across tissues. Cell 2020.

25. Harmer, D., Gilbert, M., Borman, R., Clark, K. L., Quantitative mRNA expression profiling of ACE 2, a novel homologue of angiotensin converting enzyme. FEBS Lett 2002, 532 (1-2), 107–10.

26. Kim, J.-M., Kim, H. M., Lee, E. J., Jo, H. J., Yoon, Y., Lee, N.-J., Son, J., Lee, Y.-J., Kim, M. S., Lee, Y.-P., Chae, S.-J., Park, K. R., Cho, S.-R., Park, S., Kim, S. J., Wang, E., Woo, S., Lim, A., Park, S.-J., Jang, J., Chung, Y.-S., Chin, B. S., Lee, J.-S., Lim, D., Han, M.-G., Yoo, C. K., Detection and Isolation of SARS-CoV-2 in Serum, Urine, and Stool Specimens of COVID-19 Patients from the Republic of Korea. Osong Public Health Res Perspect 2020, 11 (3), 112–117.

27. Jothimani, D., Venugopal, R., Abedin, M. F., Kaliamoorthy, I., Rela, M., COVID-19 and Liver. Journal of hepatology 2020.

28. Zhang, C., Shi, L., Wang, F.-S., Liver injury in COVID-19: management and challenges. The lancet Gastroenterology & hepatology 2020, 5 (5), 428–430.

29. Chan, J. F.-W., Zhang, A. J., Yuan, S., Poon, V. K.-M., Chan, C. C.-S., Lee, A. C.-Y., Chan, W.-M., Fan, Z., Tsoi, H.-W., Wen, L., Simulation of the clinical and pathological manifestations of Coronavirus Disease 2019 (COVID-19) in golden Syrian hamster model: implications for disease pathogenesis and transmissibility. Clinical Infectious Diseases 2020.

30. Vetter, P., Eberhardt, C., Meyer, B., Martinez, P., Torriani, G., Pigny, F., Lemeille, S., Cordey, S., Laubscher, F., Vu, D.-L., Daily viral kinetics and innate and adaptive immune responses assessment in COVID-19: a case series. medRxiv 2020.

31. Bastard, P., Rosen, L. B., Zhang, Q., Michailidis, E., Hoffmann, H.-H., Zhang, Y., Dorgham, K., Philippot, Q., Rosain, J., Béziat, V., Manry, J., Shaw, E., Haljasmägi, L., Peterson, P., Lorenzo, L., Bizien, L., Trouillet-Assant, S., Dobbs, K., de Jesus, A. A., Belot, A., Kallaste, A., Catherinot, E., Tandjaoui-Lambiotte, Y., Le Pen, J., Kerner, G., Bigio, B., Seeleuthner, Y., Yang, R., Bolze, A., Spaan, A. N., Delmonte, O. M., Abers, M. S., Aiuti, A., Casari, G., Lampasona, V., Piemonti, L., Ciceri, F., Bilguvar, K., Lifton, R. P., Vasse, M., Smadja, D. M., Migaud, M., Hadjadj, J., Terrier, B., Duffy, D., Quintana-Murci, L., van de Beek, D., Roussel, L., Vinh, D. C., Tangye, S. G., Haerynck, F., Dalmau, D., Martinez-Picado, J., Brodin, P., Nussenzweig, M. C., Boisson-Dupuis, S., Rodríguez-Gallego, C., Vogt, G., Mogensen, T. H., Oler, A. J., Gu, J., Burbelo, P. D., Cohen, J., Biondi, A., Bettini, L. R., D’Angio, M., Bonfanti, P., Rossignol, P., Mayaux, J., Rieux-Laucat, F., Husebye, E. S., Fusco, F., Ursini, M. V., Imberti, L., Sottini, A., Paghera, S., Quiros-Roldan, E., Rossi, C., Castagnoli, R., Montagna, D., Licari, A., Marseglia, G. L., Duval, X., Ghosn, J., Tsang, J. S., Goldbach-Mansky, R., Kisand, K., Lionakis, M. S., Puel, A., Zhang, S.-Y., Holland, S. M., Gorochov, G., Jouanguy, E., Rice, C. M., Cobat, A., Notarangelo, L. D., Abel, L., Su, H. C., Casanova, J.-L., Auto-antibodies against type I IFNs in patients with life-threatening COVID-19. Science 2020, eabd4585.

32. Zhang, Q., Bastard, P., Liu, Z., Le Pen, J., Moncada-Velez, M., Chen, J., Ogishi, M., Sabli, I. K. D., Hodeib, S., Korol, C., Rosain, J., Bilguvar, K., Ye, J., Bolze, A., Bigio, B., Yang, R., Arias, A. A., Zhou, Q., Zhang, Y., Onodi, F., Korniotis, S., Karpf, L., Philippot, Q., Chbihi, M., Bonnet-Madin, L., Dorgham, K., Smith, N., Schneider, W. M., Razooky, B. S., Hoffmann, H.-H., Michailidis, E., Moens, L., Han, J. E., Lorenzo, L., Bizien, L., Meade, P., Neehus, A.-L., Ugurbil, A. C., Corneau, A., Kerner, G., Zhang, P., Rapaport, F., Seeleuthner, Y., Manry, J., Masson, C., Schmitt, Y., Schlüter, A., Le Voyer, T., Khan, T., Li, J., Fellay, J., Roussel, L., Shahrooei, M., Alosaimi, M. F., Mansouri, D., Al-Saud, H., Al-Mulla, F., Almourfi, F., Al-Muhsen, S. Z., Alsohime, F., Al Turki, S., Hasanato, R., van de Beek, D., Biondi, A., Bettini, L. R., D’Angio, M., Bonfanti, P., Imberti, L., Sottini, A., Paghera, S., Quiros-Roldan, E., Rossi, C., Oler, A. J., Tompkins, M. F., Alba, C., Vandernoot, I., Goffard, J.-C., Smits, G., Migeotte, I., Haerynck, F., Soler-Palacin, P., Martin-Nalda, A., Colobran, R., Morange, P.-E., Keles, S., Çölkesen, F., Ozcelik, T., Yasar, K. K., Senoglu, S., Karabela, Ş.N., Gallego, C. R., Novelli, G., Hraiech, S., Tandjaoui-Lambiotte, Y., Duval, X., Laouénan, C., Snow, A. L., Dalgard, C. L., Milner, J., Vinh, D. C., Mogensen, T. H., Marr, N., Spaan, A. N., Boisson, B., Boisson-Dupuis, S., Bustamante, J., Puel, A., Ciancanelli, M., Meyts, I., Maniatis, T., Soumelis, V., Amara, A., Nussenzweig, M., García-Sastre, A., Krammer, F., Pujol, A., Duffy, D., Lifton, R., Zhang, S.-Y., Gorochov, G., Béziat, V., Jouanguy, E., Sancho-Shimizu, V., Rice, C. M., Abel, L., Notarangelo, L. D., Cobat, A., Su, H. C., Casanova, J.-L., Inborn errors of type I IFN immunity in patients with life-threatening COVID-19. Science 2020, eabd4570.

33. Hadjadj, J., Yatim, N., Barnabei, L., Corneau, A., Boussier, J., Pere, H., Charbit, B., Bondet, V., Chenevier-Gobeaux, C., Breillat, P., Impaired type I interferon activity and exacerbated inflammatory responses in severe Covid-19 patients. MedRxiv 2020.

34. Fenner, F., Bachmann, P. A., Gibbs, E. P. J., Murphy, F. A., Studdert, M. J., White, D. O., Pathogenesis: Infection and the Spread of Viruses in the Body. Veterinary Virology 1987, 133.

35. Dogra, P., Adolphi, N. L., Wang, Z., Lin, Y.-S., Butler, K. S., Durfee, P. N., Croissant, J. G., Noureddine, A., Coker, E. N., Bearer, E. L., Cristini, V., Brinker, C. J., Establishing the effects of mesoporous silica nanoparticle properties on in vivo disposition using imaging-based pharmacokinetics. Nature Communications 2018, 9 (1), 4551.

36. Dogra, P., Butner, J. D., Chuang, Y.-l., Caserta, S., Goel, S., Brinker, C. J., Cristini, V., Wang, Z., Mathematical modeling in cancer nanomedicine: a review. Biomedical Microdevices 2019, 21 (2), 40.

37. Dogra, P., Butner, J. D., Nizzero, S., Ruiz Ramirez, J., Noureddine, A., Pelaez, M. J., Elganainy, D., Yang, Z., Le, A. D., Goel, S., Leong, H. S., Koay, E. J., Brinker, C. J., Cristini, V., Wang, Z., Image-guided mathematical modeling for pharmacological evaluation of nanomaterials and monoclonal antibodies. Wiley Interdiscip Rev Nanomed Nanobiotechnol 2020, e1628.

38. Dogra, P., Butner, J. D., Ramírez, J. R., Chuang, Y.-l., Noureddine, A., Brinker, C. J., Cristini, V., Wang, Z., A mathematical model to predict nanomedicine pharmacokinetics and tumor delivery. Computational and Structural Biotechnology Journal 2020, 18, 518–531.

39. Goel, S., Ferreira, C. A., Dogra, P., Yu, B., Kutyreff, C. J., Siamof, C. M., Engle, J. W., Barnhart, T. E., Cristini, V., Wang, Z., Size-Optimized Ultrasmall Porous Silica Nanoparticles Depict Vasculature-Based Differential Targeting in Triple Negative Breast Cancer. Small 2019.

40. Goel, S., Zhang, G., Dogra, P., Nizzero, S., Cristini, V., Wang, Z., Hu, Z., Li, Z., Liu, X., Shen, H., Sequential deconstruction of composite drug transport in metastatic breast cancer. Science Advances 2020, 6 (26), eaba4498.

41. Sarin, H., Physiologic upper limits of pore size of different blood capillary types and another perspective on the dual pore theory of microvascular permeability. Journal of angiogenesis research 2010, 2 (1), 14.

42. Qin, J., You, C., Lin, Q., Hu, T., Yu, S., Zhou, X.-H., Estimation of incubation period distribution of COVID-19 using disease onset forward time: a novel cross-sectional and forward follow-up study. medRxiv 2020.

43. Zheng, S., Fan, J., Yu, F., Feng, B., Lou, B., Zou, Q., Xie, G., Lin, S., Wang, R., Yang, X., Viral load dynamics and disease severity in patients infected with SARS-CoV-2 in Zhejiang province, China, January-March 2020: retrospective cohort study. bmj 2020, 369.

44. Iyer, A. S., Jones, F. K., Nodoushani, A., Kelly, M., Becker, M., Slater, D., Mills, R., Teng, E., Kamruzzaman, M., Garcia-Beltran, W. F., Persistence and decay of human antibody responses to the receptor binding domain of SARS-CoV-2 spike protein in COVID-19 patients. Science Immunology 2020, 5 (52).

45. Isho, B., Abe, K. T., Zuo, M., Jamal, A. J., Rathod, B., Wang, J. H., Li, Z., Chao, G., Rojas, O. L., Bang, Y. M., Persistence of serum and saliva antibody responses to SARS-CoV-2 spike antigens in COVID-19 patients. Science Immunology 2020, 5 (52).

46. Tillett, R. L., Sevinsky, J. R., Hartley, P. D., Kerwin, H., Crawford, N., Gorzalski, A., Laverdure, C., Verma, S. C., Rossetto, C. C., Jackson, D., Genomic evidence for reinfection with SARS-CoV-2: a case study. The Lancet Infectious Diseases 2020.

47. Seow, J., Graham, C., Merrick, B., Acors, S., Pickering, S., Steel, K. J. A., Hemmings, O., O’Byrne, A., Kouphou, N., Galao, R. P., Betancor, G., Wilson, H. D., Signell, A. W., Winstone, H., Kerridge, C., Huettner, I., Jimenez-Guardeño, J. M., Lista, M. J., Temperton, N., Snell, L. B., Bisnauthsing, K., Moore, A., Green, A., Martinez, L., Stokes, B., Honey, J., Izquierdo-Barras, A., Arbane, G., Patel, A., Tan, M. K. I., O’Connell, L., O’Hara, G., MacMahon, E., Douthwaite, S., Nebbia, G., Batra, R., Martinez-Nunez, R., Shankar-Hari, M., Edgeworth, J. D., Neil, S. J. D., Malim, M. H., Doores, K. J., Longitudinal observation and decline of neutralizing antibody responses in the three months following SARS-CoV-2 infection in humans. Nature Microbiology 2020.

48. Gupta, A., Madhavan, M. V., Sehgal, K., Nair, N., Mahajan, S., Sehrawat, T. S., Bikdeli, B., Ahluwalia, N., Ausiello, J. C., Wan, E. Y., Extrapulmonary manifestations of COVID-19. Nature medicine 2020, 26 (7), 1017–1032.

49. Cai, X., Ma, Y., Li, S., Chen, Y., Rong, Z., Li, W., Clinical Characteristics of 5 COVID-19 Cases With Non-respiratory Symptoms as the First Manifestation in Children. Frontiers in Pediatrics 2020, 8, 258.

50. Liao, M., Liu, Y., Yuan, J., Wen, Y., Xu, G., Zhao, J., Chen, L., Li, J., Wang, X., Wang, F., The landscape of lung bronchoalveolar immune cells in COVID-19 revealed by single-cell RNA sequencing. MedRxiv 2020.

51. Perea, L., Rodrigo-Troyano, A., Cantó, E., Domínguez-Á;lvarez, M., Giner, J., Sanchez-Reus, F., Villar-García, J., Quero, S., García-Núñez, M., Marín, A., Reduced airway levels of fatty-acid binding protein 4 in COPD: relationship with airway infection and disease severity. Respiratory Research 2020, 21 (1), 1–8.

52. Chen, Y., Feng, Z., Diao, B., Wang, R., Wang, G., Wang, C., Tan, Y., Yuan, Z., The novel severe acute respiratory syndrome coronavirus 2 (SARS-CoV-2) directly decimates human spleens and lymph nodes. medRxiv 2020.03. 27.20045427. 2020.

53. Sanders, J. M., Monogue, M. L., Jodlowski, T. Z., Cutrell, J. B., Pharmacologic treatments for coronavirus disease 2019 (COVID-19): a review. Jama 2020, 323 (18), 1824–1836.

54. Hung, I. F.-N., Lung, K.-C., Tso, E. Y.-K., Liu, R., Chung, T. W.-H., Chu, M.-Y., Ng, Y.-Y., Lo, J., Chan, J., Tam, A. R., Triple combination of interferon beta-1b, lopinavir–ritonavir, and ribavirin in the treatment of patients admitted to hospital with COVID-19: an open-label, randomised, phase 2 trial. The Lancet 2020, 395 (10238), 1695–1704.

55. Zhou, Q., Wei, X.-S., Xiang, X., Wang, X., Wang, Z.-H., Chen, V., Shannon, C. P., Tebbutt, S. J., Kollmann, T. R., Fish, E. N., Interferon-a2b treatment for COVID-19. MedRxiv 2020.

56. Beigel, J. H., Tomashek, K. M., Dodd, L. E., Mehta, A. K., Zingman, B. S., Kalil, A. C., Hohmann, E., Chu, H. Y., Luetkemeyer, A., Kline, S., Lopez de Castilla, D., Finberg, R. W., Dierberg, K., Tapson, V., Hsieh, L., Patterson, T. F., Paredes, R., Sweeney, D. A., Short, W. R., Touloumi, G., Lye, D. C., Ohmagari, N., Oh, M.-d., Ruiz-Palacios, G. M., Benfield, T., Fätkenheuer, G., Kortepeter, M. G., Atmar, R. L., Creech, C. B., Lundgren, J., Babiker, A. G., Pett, S., Neaton, J. D., Burgess, T. H., Bonnett, T., Green, M., Makowski, M., Osinusi, A., Nayak, S., Lane, H. C., Remdesivir for the Treatment of Covid-19 — Final Report. New England Journal of Medicine 2020.

57. Hu, X., Shang, S., Nestorov, I., Hasan, J., Seddighzadeh, A., Dawson, K., Sperling, B., Werneburg, B., COMPARE: Pharmacokinetic profiles of subcutaneous peginterferon beta-1a and subcutaneous interferon beta-1a over 2 weeks in healthy subjects. Br J Clin Pharmacol 2016, 82 (2), 380–388.

58. Bar-On, Y. M., Flamholz, A., Phillips, R., Milo, R., SARS-CoV-2 (COVID-19) by the numbers. Elife 2020, 9, e57309.

59. Pezzini, A., Padovani, A., Lifting the mask on neurological manifestations of COVID-19. Nature Reviews Neurology 2020, 1–9.

60. Wang, Z., Bordas, V., Deisboeck, T. S., Identification of Critical Molecular Components in a Multiscale Cancer Model Based on the Integration of Monte Carlo, Resampling, and ANOVA. Front Physiol 2011, 2, 35–35.

61. Wang, Z., Deisboeck, T. S., Cristini, V., Development of a sampling-based global sensitivity analysis workflow for multiscale computational cancer models. IET systems biology 2014, 8 (5), 191–7.

62. De Boer, R. J., Perelson, A. S., Quantifying T lymphocyte turnover. Journal of theoretical biology 2013, 327, 45–87.

63. Westera, L., Drylewicz, J., den Braber, I., Mugwagwa, T., van der Maas, I., Kwast, L., Volman, T., van de Weg-Schrijver, E. H., Bartha, I., Spierenburg, G., Gaiser, K., Ackermans, M. T., Asquith, B., de Boer, R. J., Tesselaar, K., Borghans, J. A., Closing the gap between T-cell life span estimates from stable isotope-labeling studies in mice and humans. Blood 2013, 122 (13), 2205–12.

64. Auner, H. W., Beham-Schmid, C., Dillon, N., Sabbattini, P., The life span of short-lived plasma cells is partly determined by a block on activation of apoptotic caspases acting in combination with endoplasmic reticulum stress. Blood 2010, 116 (18), 3445–3455.

65. Andraud, M., Lejeune, O., Musoro, J. Z., Ogunjimi, B., Beutels, P., Hens, N., Living on three time scales: the dynamics of plasma cell and antibody populations illustrated for hepatitis a virus. PLoS Comput Biol 2012, 8 (3), e1002418–e1002418.

66. Bortnick, A., Allman, D., What is and what should always have been: long-lived plasma cells induced by T cell-independent antigens. J Immunol 2013, 190 (12), 5913–5918.

67. Kumar, B. V., Connors, T. J., Farber, D. L., Human T Cell Development, Localization, and Function throughout Life. Immunity 2018, 48 (2), 202–213.

68. Collin, M., Bigley, V., Human dendritic cell subsets: an update. Immunology 2018, 154 (1), 3–20.

69. Gray, D., Lifespan of Immune Cells and Molecules. In Encyclopedia of Immunology (Second Edition), Delves, P. J., Ed. Elsevier: Oxford, 1998; pp 1579–1583.

70. Alloatti, A., Kotsias, F., Magalhaes, J. G., Amigorena, S., Dendritic cell maturation and cross-presentation: timing matters! Immunol Rev 2016, 272 (1), 97–108.

71. Nair, S., Archer, G. E., Tedder, T. F., Isolation and generation of human dendritic cells. Curr Protoc Immunol 2012, Chapter 7, Unit7.32-Unit7.32.

72. Bianconi, E., Piovesan, A., Facchin, F., Beraudi, A., Casadei, R., Frabetti, F., Vitale, L., Pelleri, M. C., Tassani, S., Piva, F., Perez-Amodio, S., Strippoli, P., Canaider, S., An estimation of the number of cells in the human body. Ann Hum Biol 2013, 40 (6), 463–71.

73. Bittersohl, H., Steimer, W., Chapter 9 - Intracellular concentrations of immunosuppressants. In Personalized Immunosuppression in Transplantation, Oellerich, M., Dasgupta, A., Eds. Elsevier: San Diego, 2016; pp 199–226.

74. Alberts, B., Johnson, A., Lewis, J., Raff, M., Roberts, K., Walter, P., Molecular Biology of the Cell 4th ed. New York: Garland Science; 2002.

75. Ruedl, C., Koebel, P., Bachmann, M., Hess, M., Karjalainen, K., Anatomical Origin of Dendritic Cells Determines Their Life Span in Peripheral Lymph Nodes. The Journal of Immunology 2000, 165 (9), 4910–4916.

76. Macallan, D. C., Wallace, D. L., Zhang, Y., Ghattas, H., Asquith, B., de Lara, C., Worth, A., Panayiotakopoulos, G., Griffin, G. E., Tough, D. F., Beverley, P. C. L., B-cell kinetics in humans: rapid turnover of peripheral blood memory cells. Blood 2005, 105 (9), 3633–3640.

77. Benson, N., de Jongh, J., Duckworth, J. D., Jones, H. M., Pertinez, H. E., Rawal, J. K., van Steeg, T. J., Van der Graaf, P. H., Pharmacokinetic-pharmacodynamic modeling of alpha interferon response induced by a Toll-like 7 receptor agonist in mice. Antimicrobial agents and chemotherapy 2010, 54 (3), 1179–1185.

78. Ziegler, C. G. K., Allon, S. J., Nyquist, S. K., Mbano, I. M., Miao, V. N., Tzouanas, C. N., Cao, Y., Yousif, A. S., Bals, J., Hauser, B. M., Feldman, J., Muus, C., Wadsworth, M. H., Kazer, S. W., Hughes, T. K., Doran, B., Gatter, G. J., Vukovic, M., Taliaferro, F., Mead, B. E., Guo, Z., Wang, J. P., Gras, D., Plaisant, M., Ansari, M., Angelidis, I., Adler, H., Sucre, J. M. S., Taylor, C. J., Lin, B., Waghray, A., Mitsialis, V., Dwyer, D. F., Buchheit, K. M., Boyce, J. A., Barrett, N. A., Laidlaw, T. M., Carroll, S. L., Colonna, L., Tkachev, V., Peterson, C. W., Yu, A., Zheng, H. B., Gideon, H. P., Winchell, C. G., Lin, P. L., Bingle, C. D., Snapper, S. B., Kropski, J. A., Theis, F. J., Schiller, H. B., Zaragosi, L.-E., Barbry, P., Leslie, A., Kiem, H.-P., Flynn, J. L., Fortune, S. M., Berger, B., Finberg, R. W., Kean, L. S., Garber, M., Schmidt, A. G., Lingwood, D., Shalek, A. K., Ordovas-Montanes, J., Banovich, N., Barbry, P., Brazma, A., Desai, T., Duong, T. E., Eickelberg, O., Falk, C., Farzan, M., Glass, I., Haniffa, M., Horvath, P., Hung, D., Kaminski, N., Krasnow, M., Kropski, J. A., Kuhnemund, M., Lafyatis, R., Lee, H., Leroy, S., Linnarson, S., Lundeberg, J., Meyer, K., Misharin, A., Nawijn, M., Nikolic, M. Z., Ordovas-Montanes, J., Pe’er, D., Powell, J., Quake, S., Rajagopal, J., Tata, P. R., Rawlins, E. L., Regev, A., Reyfman, P. A., Rojas, M., Rosen, O., Saeb-Parsy, K., Samakovlis, C., Schiller, H., Schultze, J. L., Seibold, M. A., Shalek, A. K., Shepherd, D., Spence, J., Spira, A., Sun, X., Teichmann, S., Theis, F., Tsankov, A., van den Berge, M., von Papen, M., Whitsett, J., Xavier, R., Xu, Y., Zaragosi, L.-E., Zhang, K., SARS-CoV-2 Receptor ACE2 Is an Interferon-Stimulated Gene in Human Airway Epithelial Cells and Is Detected in Specific Cell Subsets across Tissues. Cell 2020, 181 (5), 1016-1035.e19.

79. Hou, Y. J., Okuda, K., Edwards, C. E., Martinez, D. R., Asakura, T., Dinnon, K. H., 3rd; Kato, T., Lee, R. E., Yount, B. L., Mascenik, T. M., Chen, G., Olivier, K. N., Ghio, A., Tse, L. V., Leist, S. R., Gralinski, L. E., Schäfer, A., Dang, H., Gilmore, R., Nakano, S., Sun, L., Fulcher, M. L., Livraghi-Butrico, A., Nicely, N. I., Cameron, M., Cameron, C., Kelvin, D. J., de Silva, A., Margolis, D. M., Markmann, A., Bartelt, L., Zumwalt, R., Martinez, F. J., Salvatore, S. P., Borczuk, A., Tata, P. R., Sontake, V., Kimple, A., Jaspers, I., O’Neal, W. K., Randell, S. H., Boucher, R. C., Baric, R. S., SARS-CoV-2 Reverse Genetics Reveals a Variable Infection Gradient in the Respiratory Tract. Cell 2020, 182 (2), 429-446.e14.

80. Lee, J. J., Kopetz, S., Vilar, E., Shen, J. P., Chen, K., Maitra, A., Relative Abundance of SARS-CoV-2 Entry Genes in the Enterocytes of the Lower Gastrointestinal Tract. Genes (Basel) 2020, 11 (6), 645.

81. Qi, F., Qian, S., Zhang, S., Zhang, Z., Single cell RNA sequencing of 13 human tissues identify cell types and receptors of human coronaviruses. Biochem Biophys Res Commun 2020, 526 (1), 135–140.

82. Chai, X., Hu, L., Zhang, Y., Han, W., Lu, Z., Ke, A., Zhou, J., Shi, G., Fang, N., Fan, J., Cai, J., Fan, J., Lan, F., Specific ACE2 Expression in Cholangiocytes May Cause Liver Damage After 2019-nCoV Infection. 2020.

83. Tabibian, J. H., LaRusso, N. F., Liver and Bile. In Reference Module in Biomedical Sciences, Elsevier: 2014.

84. Radisic, M., Park, H., Vunjak-Novakovic, G., Chapter Thirty-Eight - Cardiac-Tissue Engineering. In Principles of Tissue Engineering (Third Edition), Lanza, R., Langer, R., Vacanti, J., Eds. Academic Press: Burlington, 2007; pp 551–567.

85. Guo, J., Wei, X., Li, Q., Li, L., Yang, Z., Shi, Y., Qin, Y., Zhang, X., Wang, X., Zhi, X., Meng, D., Single-cell RNA analysis on ACE2 expression provides insights into SARS-CoV-2 potential entry into the bloodstream and heart injury. J Cell Physiol 2020, 235 (12), 9884–9894.

86. Lin, W., Hu, L., Zhang, Y., Ooi, J. D., Meng, T., Jin, P., Ding, X., Peng, L., Song, L., Xiao, Z., Ao, X., Xiao, X., Zhou, Q., Xiao, P., Fan, J., Zhong, Y., Single-cell Analysis of ACE2 Expression in Human Kidneys and Bladders Reveals a Potential Route of 2019-nCoV Infection. 2020.

87. Chen, R., Wang, K., Yu, J., Howard, D., French, L., Chen, Z., Wen, C., Xu, Z., The spatial and cell-type distribution of SARS-CoV-2 receptor ACE2 in human and mouse brain. 2020.

88. Valério-Gomes, B., Guimarães, D. M., Szczupak, D., Lent, R., The Absolute Number of Oligodendrocytes in the Adult Mouse Brain. Frontiers in Neuroanatomy 2018, 12, 90.

89. Gabrielsson, J., Weiner, D., Pharmacokinetic and pharmacodynamic data analysis: concepts and applications. CRC Press: 2001.

