## Supplementary material for "Innate immunity plays a key role in controlling viral load in COVID-19: mechanistic insights from a whole-body infection dynamics model": SI figure

**Short title:** A multiscale COVID-19 model
**Keywords:** COVID-19, SARS-CoV-2, mathematical modeling, viral dynamics, pharmacokinetics, in silico clinical trials

*Correspondence should be sent to:
Zhihui Wang, PhD

Or,
Prashant Dogra, PhD


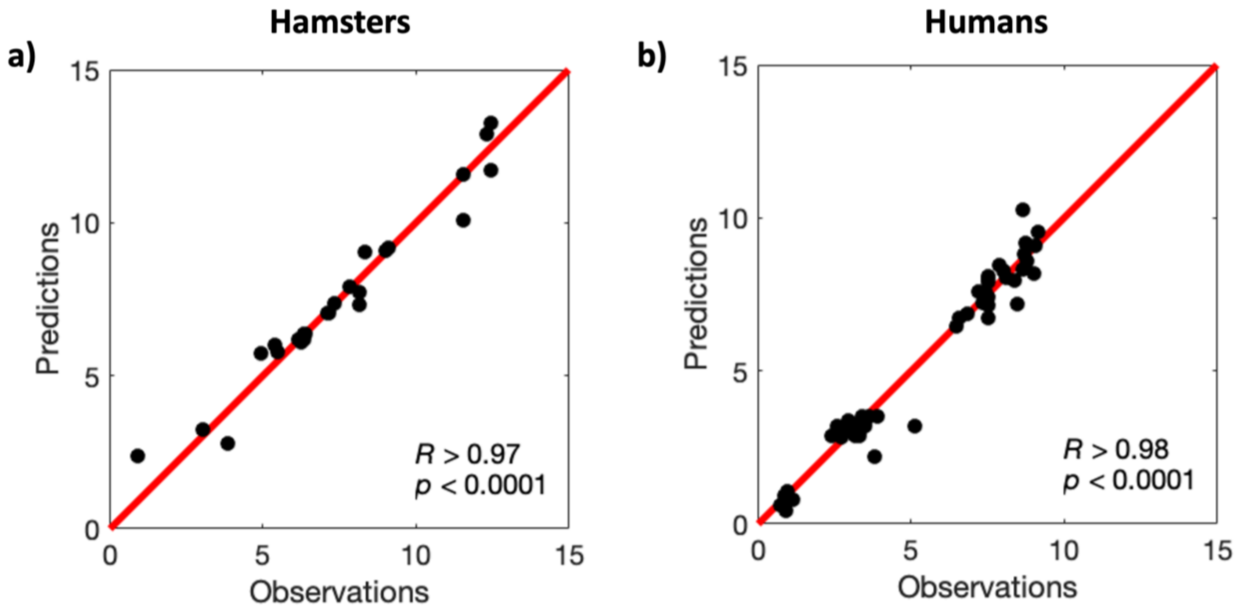


**Figure S1. Pearson correlation of model predictions to experimental data for a) hamsters and b) humans.**
